# Talin1 dysfunction is genetically linked to systemic capillary leak syndrome

**DOI:** 10.1101/2022.10.17.22280833

**Authors:** Naama Elefant, Pinelopi A Nikolopoulou, V Vassiliki Papadaki, Danit Oz-Levi, Georgia Rouni, Racheli Sion-Sarid, William JS Edwards, Christina Arapatzi, Shira Yanovsky-Dagan, Alana R Cowell, Vardiella Meiner, Vladimir Vainstein, Sofia Grammenoudi, Doron Lancet, Benjamin T Goult, Tamar Harel, Vassiliki Kostourou

## Abstract

Systemic capillary leak syndrome (SCLS) is a rare life-threatening disorder due to profound vascular leak. The trigger and the cause of the disease is currently unknown and there is no specific treatment. Here, we identified a rare heterozygous splice-site variant in the *TLN1* gene in a familial SCLS case, suggestive of autosomal dominant inheritance with incomplete penetrance. Talin1 has a key role in cell adhesions by activating and linking integrins to the actin cytoskeleton. This variant causes in-frame skipping of exon 54 and is predicted to affect talin’s c-terminal actin binding site (ABS3). Modelling the SCLS-*TLN1* variant by mimicking the actin-binding disruption in *TLN1* heterozygous endothelial cells resulted in disorganized endothelial adherens junctions. Mechanistically, we established that disruption of talin’s ABS3 sequestrates talin’s interacting partner, vinculin, at cell-extracellular matrix adhesions, leading to destabilization of the endothelial barrier. We propose that pathogenic variant in *TLN1* underlie SCLS, providing insight into the molecular mechanism of the disease which can be explored for future therapeutic interventions.

**SUMMARY:** Systemic capillary leak syndrome (SCLS) is a rare potentially lethal disease with unknown etiology and non-specific treatment. Here, we established a heterozygous splice variant of talin1, a key cell adhesion protein, as a genetic link to a familial SCLS case.

## INTRODUCTION

Systemic capillary leak syndrome (SCLS) is a rare and life-threating condition that was first described in 1960 by Clarkson and is also referred to as Clarkson’s disease (Clarkson et al., 1960). It is characterized by recurrent, transient, and spontaneous episodes of massive shifts of intravascular fluids into peripheral tissues and presents with hypovolemic shock, hemoconcentration and hypoproteinemia (Marks and Shuster, 1973). Only a few hundred sporadic cases are described world-wide with presenting ages spanning from early childhood to adulthood. Frequency and severity of episodes vary between patients. Episodes are frequently preceded by viral infections and about half of the patients report preceding symptoms including oligo-anuria, fatigue, edema, syncopal episodes, abdominal pain, nausea, myalgias, polydipsia, and a sudden increase in body weight (Amoura et al., 1997; Eo et al., 2018). Episodes are usually treated in the setting of an emergency department or an intensive care unit (ICU), with massive infusions of intravenous (IV) saline. There are no established acute pharmacologic therapies for SCLS. In several reports, the use of intravenous immunoglobulins (IVIG), terbutaline combined with aminophylline, infliximab or anti-VEGF antibodies are described with varying degrees of success (Xie et al., 2015; Eo et al., 2018). Treatment is challenging, since the provided fluids are promptly “third-spaced” due to the vascular leakage, leading to anasarca (with relative sparing of the lungs). As a result, compartment syndrome may develop in the peripheral limbs, frequently necessitating fasciotomies. Symptoms typically resolve spontaneously after 48–96 hours without specific treatment other than intense hemodynamic support and treatment of complications.

The etiology of SCLS remains unknown with most current hypotheses presupposing exaggerated or abnormal cellular signaling, either in vascular endothelium or inflammatory cells (Druey, 2014; Xie et al., 2012). Up to 80% of adult patients with SCLS are found to have a monoclonal gammopathy of unknown significance (MGUS), but the role of this finding in the pathogenicity of the disease has not been established and is not considered essential for the diagnosis of SCLS (Xie et al., 2015). In an attempt to discover germline mutations underlying the disease several papers have conducted sequencing studies of patients with SCLS. In one publication, genome-wide single nucleotide polymorphism (SNP) analysis was conducted for 12 patients revealing several loci and SNPs associated with the disease (Xie et al., 2014). In a more recent study, exome sequencing was performed for 16 adult and pediatric patients with SCLS. In this paper several candidate genes were proposed but no uniform germline exonic variants were found (Pierce et al., 2018).

Here we report on the results of the genetic investigation of three SCLS patients and segregation analysis from an extended family pedigree suggestive of autosomal dominant inheritance with incomplete penetrance. We identified a heterozygous splice-site variant c.7188+2T>C in the *TLN1* gene affecting the C-terminal region of the Talin1 protein. Talin is a key member of the integrin-mediated cell-extracellular matrix (ECM) adhesions with important functions in integrin activation, mechanotransduction, cell attachment and spreading (Calderwood et al., 2013; Goult et al., 2018). Talin1 is a 2541 amino acid protein, that is comprised of an N-terminal head domain that binds integrins connected to a C-terminal rod region comprised of 13 alpha-helix bundles, R1-R13 culminating in a dimerisation domain (DD) helix (Gingras et al., 2008). The talin rod region contains eleven vinculin binding sites (VBS) and two actin binding sites (ABS), that can be conformationally regulated by application of cytoskeletal forces. Several studies have demonstrated a significant role for talin in physiological organ function and cancer (Haining et al., 2016; Nikolopoulou et al., 2021). In the vascular system, talin1 is essential for embryonic and postnatal blood vessel development and tumour angiogenesis (Monkley et al., 2011b; Pulous et al., 2020; Nikolopoulou et al., 2022). Recently, it was shown that endothelial-specific deletion of talin caused intestinal microvascular bleeding because of defective integrin activation (Pulous et al., 2019). In addition, studies have shown that *TLN1* was downregulated in aortic tissue of patients with aortic dissection and rare heterozygous missense variants in *TLN1* were found in individuals with familial spontaneous coronary artery dissection (SCAD) (Wei et al., 2017; Turley et al., 2019). Together these studies highlight a critical role for talin1 in vascular endothelium.

Here we report an extended pedigree with SCLS and show that heterozygous dysfunction in the C-terminal actin-binding domain (ABS3) of talin perturbs the localization of its partner vinculin to adherens junctions, leading to defective remodeling of cell-cell junctions and loss of vascular integrity. Our data indicates a genetic germline etiology and provides a mechanistic insight into the cause of SCLS that can be exploited for both diagnostic and therapeutic interventions.

## RESULTS

### Clinical Reports

The proband (Fig. 1A, Individual IV-3) presented with her first episode of systemic capillary leak syndrome (SCLS) to the ER with severe hypotension, hypovolemia and hemoconcentration and was stabilized and treated with IV fluids. A proband’s relative reported episodes of SCLS mostly in his childhood and teenage years. Another proband’s relative suffered episodes of SCLS from six months of age. In childhood, he suffered a severe attack with cerebral edema which upon resolution required rehabilitation for several months. In childhood and adolescence, he suffered monthly episodes and currently experiences an episode once every 4-6 months. Another relative died at childhood. Archived medical records describing episodes of abdominal pain, hypotension, hemoconcentration and loss of consciousness that required treatment with IV fluids, are suggestive of SCLS. In addition, several members of the extended family died at a young age, possibly having SCLS without being properly diagnosed. Detailed clinical reports are available upon request.

**Figure 1.**
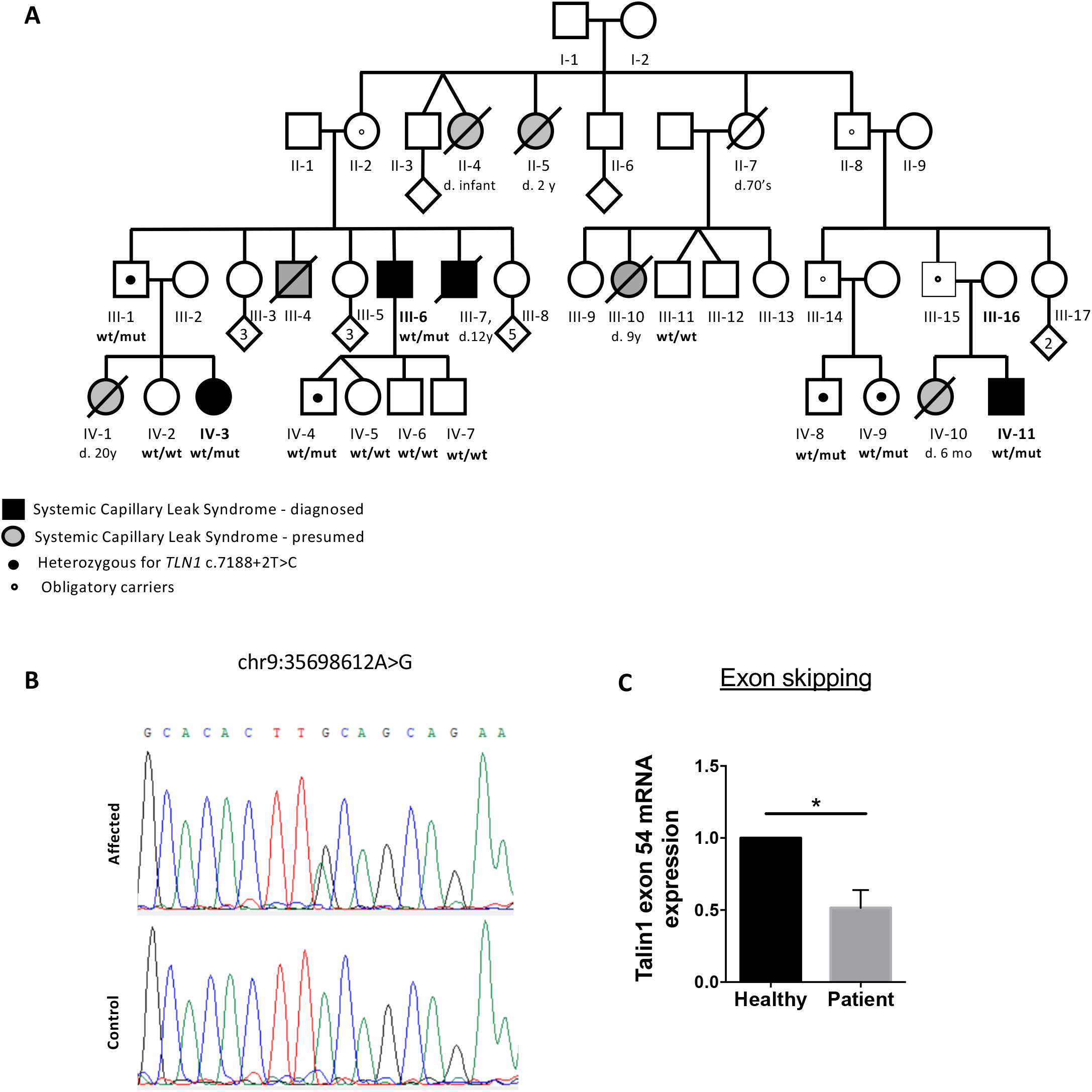
Pedigree of the familial case of SCLS and identification of SCLS-TLN1 variant. (A) Pedigree indicating affected individuals. Black-filled shapes indicate affected individuals. Grey-filled shapes indicate individuals suspected to have been affected, where DNA was not available for testing. Inheritance was presumed to be autosomal dominant with incomplete penetrance. (B) Sanger sequencing at the DNA level, showing heterozygous missense variant of slice site at exon 54 (c.7188+2T>C). (C) Quantitative Real Time PCR analysis showing the fold difference in the mRNA expression of exon 54 between control and patient fibroblasts normalized to RPLP1 reference gene.

### Identification of a heterozygous variant in *TLN1*

Exome sequencing was undertaken in all four identified patients, in search of an underlying genetic diagnosis. Following alignment to the reference genome [hg19] and variant calling, variants were filtered out if they were off-target (>8 bp from splice junction), synonymous, or had minor allele frequency (MAF) > 0.01 in the gnomAD database. A heterozygous, splice-site variant located in the intron 54 of the *TLN1* gene (chr9: g.35698612A>G[hg19]; (NM_006289.4): c.7188+2T>C) was found in all patients. This splice-site variant was not found in gnomAD nor in our in-house database of ∼13,000 exomes, and was predicted to alter splicing (SpliceAI) (Fig. 1B) (Jaganathan et al., 2019). Following this finding, segregation analysis was then preformed on several members of the extended family, and four carriers of the *TLN1* variant were identified (ages 13-62). These individuals had no history of syncope or sudden hypotension. Taken together, our genetic analysis identified a heterozygous splice variant with incomplete penetrance and variability in the age of first presentation, in this familial case of SCLS.

### The SCLS *TLN1* variant causes exon skipping at the RNA level

In order to determine whether the *TLN1* variant affects splicing, we analyzed the cDNA sequence of two patients. Two tissues were analyzed (lymphocytes and fibroblasts) to examine whether the effect of the variant was tissue specific. Reverse transcription-PCR on RNA extracted from both tissues revealed the existence of a lower molecular weight band in the affected individual in addition to the wild-type band, suggesting exon skipping (Fig. S1A). Sequencing of the lower band after gel extraction revealed skipping of exon 54 (Fig. S1B). Thus, we concluded that the *TLN1* c.7188+2T>C variant led to a heterozygous in-frame deletion of 63 nucleotides. To quantify the percentage of talin’s transcript with the exon skipping event, real time-PCR was performed on cDNA from fibroblasts derived from a SCLS patient and a healthy control relative using specific primers that targeted the affected area. The patient fibroblasts displayed a mean 50% reduction in mRNA transcript that contains exon 54, confirming the mis-splicing effect of the identified *TLN1* variant (Fig.1C).

### The SCLS *TLN1* variant results in disturbance of the 13^th^ talin rod domain, R13

The deletion of exon 54 results in the in-frame deletion of 21 amino acids from the talin primary sequence. These residues, V2376-A2396 inclusive, are situated towards the C-terminus of the talin protein in the 13^th^ rod domain, R13 (Fig. 2A and B). The structure of Talin’s R13 is a 5-helix bundle and the *TLN1* variant results in deletion of part of the 59^th^ helix of talin1 (Fig. 2B, magenta). R13 is an important domain for talin function, as it forms part of the C-terminal actin binding site, ABS3 in conjunction with the adjacent dimerization domain (DD) (Fig. 2B). Binding of actin to ABS3 has been mapped to two interfaces on the R13-DD module, one on R13 centered around a basic “KVK” surface (orange), and a second on the DD coiled coil centered around R2510 (green) (Fig. 2B). Mutations that disrupt either the KVK (K2443D/V2444D/K2445D) or R2510 (R2510A) surfaces have a significant impact on actin binding to ABS3 without perturbing structural integrity or dimerisation (Gingras et al., 2008). Helix 58 of R13 (yellow) is also a vinculin binding site (VBS). Because of the proximity of the SCLS deletion to the actin- and vinculin-binding sites in R13 we wanted to establish the impact of the SCLS deletion on the biochemical and biophysical properties of talin.

**Figure 2.**
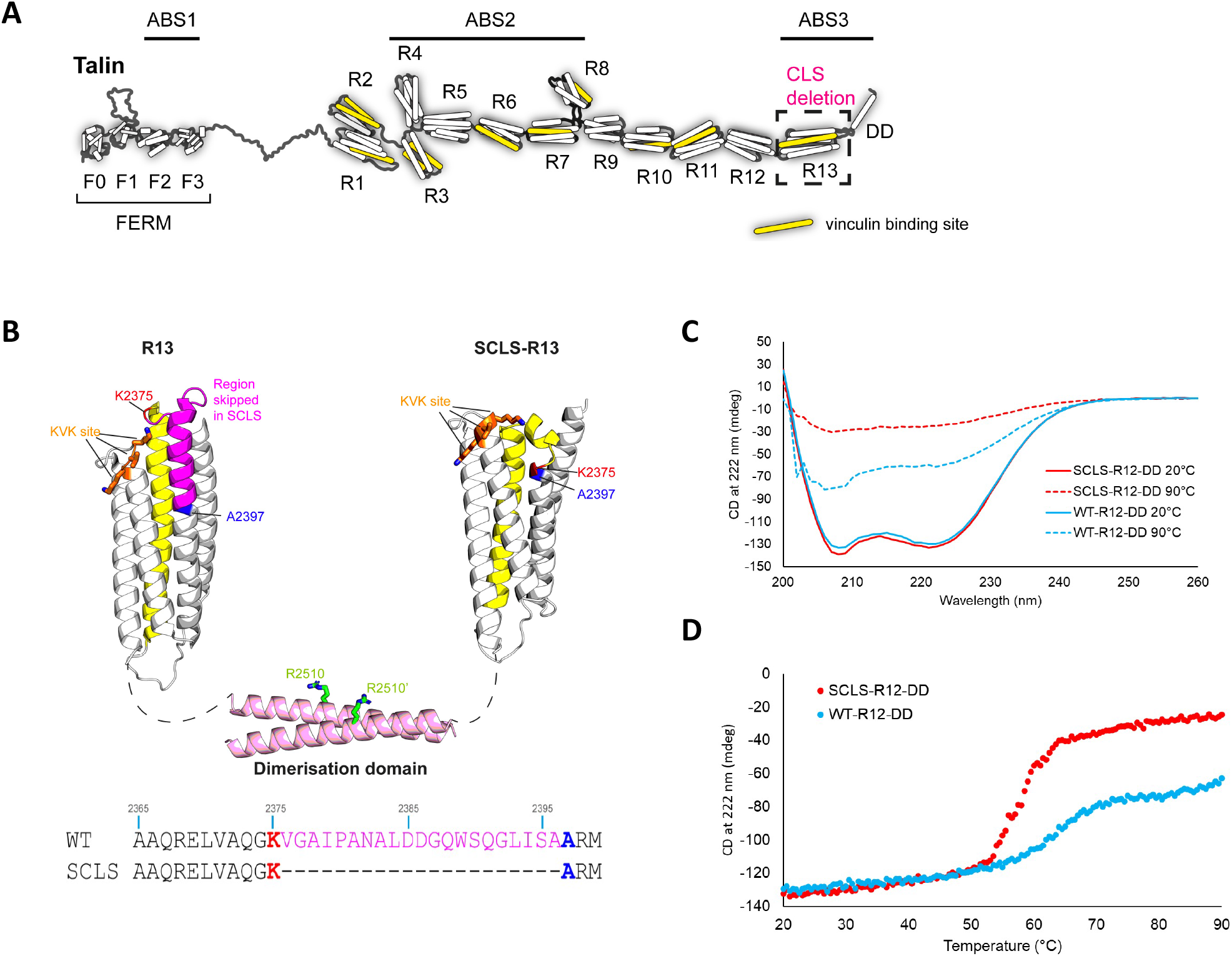
Structural analysis of the SCLS-TLN1 variant protein indicates a destabilised R13 domain. (A) The domain structure of talin contains an N-terminal FERM domain and a large rod region comprised of 13 helical bundles, R1–R13 ending in a dimerisation domain (DD). The three actin binding sites (ABS) are highlighted. The 11 vinculin binding sites (VBS) are shown in yellow. The R13 domain where the SCLS variant leads to deletion of 21 amino acids is highlighted. (B) The structure of R13 and the dimerisation domain DD (left) and the predicted structural model of SCLS-R13 (right). The KVK site (residues K2443/V2444/K2445) and the R2510 site that are required for actin binding are shown in orange and green respectively. The 21 residues skipped in SCLS are shown in magenta. K2375 (red) and A2397 (blue) are the residues immediately before and after the skipped region. To highlight the distortion the VBS helix at the back of the wild-type is shown in yellow, in the SCLS this helix is broken. Sequence alignment of part of R13 containing the skipped exon. Wild-type (top), and SCLS (bottom). (C-D) Circular dichroism (CD) analysis. (C) Far-UV spectral analysis (between 200-260 nm wavelengths) of 0.5 mg/ml WT-R12-DD (blue) and SCLS-R12-DD (red) at 20°C and at 90°C. (D) Melting curves at 222 nm for 0.5 mg/ml (blue) wild-type WT-R12-DD and (red) SCLS-R12-DD measuring thermal stability over increasing temperature from 20-90°C.

To visualize the effects of the exon skipping on the structural integrity of R13, we first generated a structural model of SCLS-R13 using the protein prediction software AlphaFold2 (Jumper et al., 2021). Comparison of the structure of WT-R13 with the structural model of SCLS-R13 revealed that the SCLS deletion severely distorts the R13 structure (Fig. 2B) and introduces a kink into the region of R13 known to be important for actin-binding.

We next generated expression constructs of ABS3 domain (R13-DD) and R12-R13-DD (R12-DD), both wildtype and with the SCLS deletion and attempted to express and purify them from E. coli using standard protocols. As previously, the wildtype R13-DD and R12-DD expressed and purified nicely, however the SCLS-R13-DD was insoluble, precluding it from further study. The SCLS-R12-DD behaved slightly better and was able to be studied. Far-UV circular dichroism (CD) analysis supported the modelling as both WT-R12-DD and SCLS-R12-DD have very similar CD spectra at 20°C with the characteristic double minima peaks at 208 and 222 nm indicative of both being predominantly alpha-helical (Fig. 2C). R13 is the most stable domain in talin unfolding at >90°C and this stability is evident in the thermal melting curves of the WT-R12-DD where two unfolding events are evident, one at ∼60°C which is the R12 unfolding, and then the start of a second unfolding event at 90°C (Fig. 2D). Even at 90°C, WT R13 is still mostly folded which is also seen in the CD spectra (Fig. 2C). In contrast, the R12-DD SCLS showed a single unfolding event at 59°C after which the whole protein is fully unfolded (Fig. 2D). Together this biophysical analysis confirms that the SCLS deletion destabilizes the R13 domain.

We next wanted to study the binding of WT-R12-DD and SCLS-R12-DD constructs to actin using the well-established actin co-sedimentation assay. However, while the WT-R12-DD remained entirely in the supernatant when centrifuged at high speeds, the SCLS-R12-DD pellets by itself (Fig. S2A), indicating the deletion renders the protein only sparingly soluble and aggregation prone and not suitable for cosedimentation based assays.

Finally, we assessed the ability of the SCLS-R12-DD to bind to the vinculin. Whereas, actin-binding to talin involves a surface of the folded domains engaging to the actin filament, vinculin binding requires the talin rod domain to unfold and expose the VBS to enable binding, including the one located at R13 (Papagrigoriou et al., 2004). Given that the SCLS deletion destabilizes the R13, we examined how this impacted vinculin binding to R13, by performing size exclusion chromatography (SEC) using talin and the talin-binding Vd1 domain (residues 1–258) of vinculin. Despite being 21 amino acids smaller than the wildtype the SCLS-R12-DD elutes slightly earlier suggesting that it is less compact in structure (Fig. S2B). The SEC experiment in the presence of a 1:1 ratio of talin proteins and vinculin Vd1 domain showed that both the wildtype R12-DD and SCLS-R12-DD bound equally well to the Vd1 at 25°C (Fig. S2B).

In summary, the result of the exon skipping at the protein level substantially disrupts the talin R13 domain which impacts its stability and we predict its ability to bind actin, however vinculin binding to R13 is not noticeably affected.

### The SCLS *TLN1* variant does not affect fibroblast adhesion properties

Mutations at splice sites often lead to decreased protein expression (Maquat and Carmichael, 2001). To examine this, we performed western blot and immunofluorescence analysis in skin fibroblasts derived from the patient IV-11 and an unaffected healthy relative control (individual III-16). We could not detect any alteration in talin1 protein expression or localization at adhesion sites in patient and control fibroblasts (Fig. S3A and B). Since fibroblasts also express the closely related *TLN2* gene (Zhang et al., 2008), we investigated whether the talin2 protein expression is altered in compensation to the SCLS-*TLN1* variant. Similar to talin1, talin2 expression and localisation at adhesion sites was unaffected in patient and control fibroblasts (Fig. S3A and B).

As talin is a critical regulator of cell-ECM adhesions (Liu et al., 2015; Monkley et al., 2011b), we investigated the effect of the SCLS-*TLN1* variant in cell-ECM adhesion formation and signaling. Immunofluorescence and western blot analysis in patient and control fibroblasts revealed that the SCLS-*TLN1* variant does not change the expression nor the localization of key adhesion proteins, including paxillin and vinculin, and does not affect the number nor the size of cell-ECM adhesions (Fig. 3A and B).

**Figure 3.**
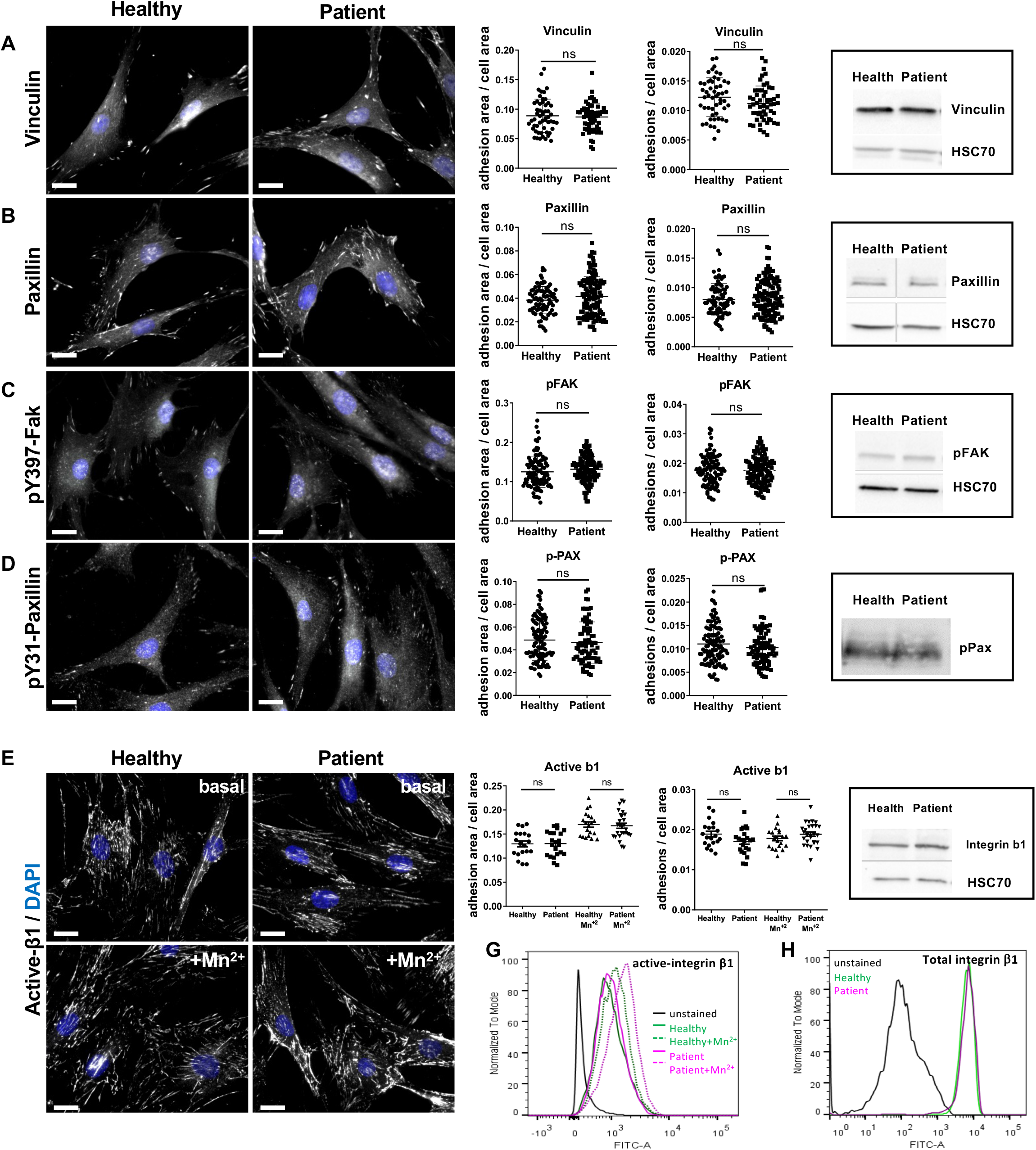
The SCLS-TLN1 variant does not affect the adhesion properties of fibroblasts. Immunofluorescent staining of SCLS patient and healthy donor fibroblasts for (A) vinculin, (B) total paxillin, (C) pY397-FAK, (D) pY118-paxillin and (E) active integrin -β1 in the absence (basal) or presence (Mn2+) of MnCl2 were used to assess cell-ECM adhesions. Nuclei were stained with DAPI. Scale bars A-D, 15μm; E, 20μm. Graphs display the quantification of the cell-ECM adhesion area (μm2) and the number of cell-ECM adhesion sites per cell area (μm2) measured with IMARIS software. Data is grouped from at least 3 independent experiments, n of cells; vinculin: Healthy=54, Patient=58; total-Paxillin: Healthy=82, Patient=123; pY397-FAK: Healthy=98, Patient=112; pY31-Paxillin: Healthy=111, Patient=88; basal active integrin b1: Healthy=67, Patient=52; induced active integrin β1: Healthy=61, Patient=74. Statistical analysis, Mann–Whitney rank-sum test. ns, no statistically significant difference. Representative western blots of cell-ECM adhesion protein expression in lysates from healthy and patient fibroblasts. HSC-70 acts as the loading control. (F) FACS histograms of patient and control fibroblasts incubated with or without Mn^2+^ and immunostained with the activation epitope-reporting 9EG7 antibody. (G) FACs histograms of total integrin-β1 cell surface levels in patient and healthy donor fibroblasts

Additional analysis of cell-ECM adhesion signaling revealed similar activation of pY397-FAK and pY31-Paxillin, in patient and control skin fibroblasts (Fig. 3C, D). In accordance with these findings, activation of major downstream signalling pathways, including pY44/42-MAPK and p-Akt were not affected (Fig. S4A and B).

Given the established role of talin in integrin activation (Calderwood et al., 2013), we examined whether the SCLS-*TLN1* variant alters integrin activation in patient and healthy donor skin fibroblasts. Immunofluorescent staining and flow cytometry analysis using an activation-epitope reporting antibody of integrin β1-subunit, showed that both basal and Mn^2+^-induced activation of integrin β1 was preserved in patient fibroblasts (Fig. 3E, F). In addition, total levels of β1-integrin expression (Fig. 3E) and surface availability, as determined by flow cytometry (Fig. 3G), remained unchanged in patient and control fibroblasts. Taken together these data indicate that the SCLS-*TLN1* variant does not affect integrin activation.

To study the functionality of cell-ECM adhesions, we performed live-cell imaging and confirmed that the SCLS-*TLN1* variant does not impair cell adhesion nor spreading on different substrates including plastic, collagen and fibronectin (Fig. S5A-C and video 1).

### Platelet function is unaffected in SCLS patient

Previous studies have shown that disruption of *TLN1* in platelets causes defects in platelet aggregation, thrombus formation and extensive bleeding in mice (Nieswandt et al., 2007; Petrich et al., 2007). Specifically, the Talin-induced integrin activation has been shown to be required for fibrin clotting (Monkley et al., 2011a). Platelet dysfunction can lead to prolonged bleeding which might contribute to SCLS symptoms. To examine whether the SCLS-*TLN1* variant affects the patient’s normal platelet activity we performed ex-vivo platelet adhesion studies. Platelets derived from both a SCLS patient and an unaffected relative donor had normal size and displayed normal aggregation to the surface (Fig. S6). Additionally, none of the patients showed any signs of aberrant clotting in their medical history or clinical examination during an attack. Together, this suggests that platelet function is not overly affected by the SCLS-*TLN1* variant.

### Generation of an *in vitro* SCLS-mimicking model in endothelial cells

Given the normal adhesion capability of patient fibroblasts and the characteristic vascular leakiness associated with the disease, we hypothesized that the endothelial cells which form blood vessels, are the key cellular player in SCLS. Thus, we set out to model the effect of the SCLS-*TLN1* variant in primary mouse endothelial cells. We have shown previously that endothelial cells express only Talin1, and that Talin2 is not expressed upon deletion of Talin1 as a compensatory mechanism (Monkley et al., 2011b). Given that the SCLS-*TLN1* variant disrupts ABS3, we generated an *in vitro* cellular model of SCLS, mimicking the disruption in actin binding region. We showed previously that an R2510A point mutation in the ABS3 disrupts talin binding to actin without effecting talin dimerization (Gingras et al., 2008) and so we used this mutant to specifically disrupt ABS3 actin binding. To model the haploinsufficiency of the SCLS mutation found in the patients, we used mouse primary endothelial cells isolated from an inducible endothelial-specific talin1 mouse model (*pdgfBB*iCreER^T2^-Talin1 flox/+). We transfected these endothelial cells with GFP-Talin1 expression constructs carrying R2510A mutation (SCLS EC-Tln^mut^) or wild-type protein (control EC-Tln^wt^). Tamoxifen-induced Cre activation *in vitro* results in genetic disruption of only one talin1 allele and thus ensures expression of the SCLS-mimicking talin1 R2510A mutant protein or control wild-type talin1, in a talin1 heterozygous background. Both constructs were fused to GFP to enable visualization of the expressed proteins. Both the SCLS-talin1 mutant and the control talin1 proteins were localized at cell-ECM adhesions (Fig. 4A). To validate further this experimental set up, we examined whether SCLS-talin1 mutation affects integrin activation. In agreement to the findings in patient fibroblasts, immunofluorescence analysis of active integrin β1 showed no difference between SCLS-talin1 mutant and control endothelial cells (Fig. 4B and C). Taken together, this experimental set up enables us to specifically analyze defective talin ABS3 binding in a physiologically relevant, heterozygous system.

**Figure 4.**
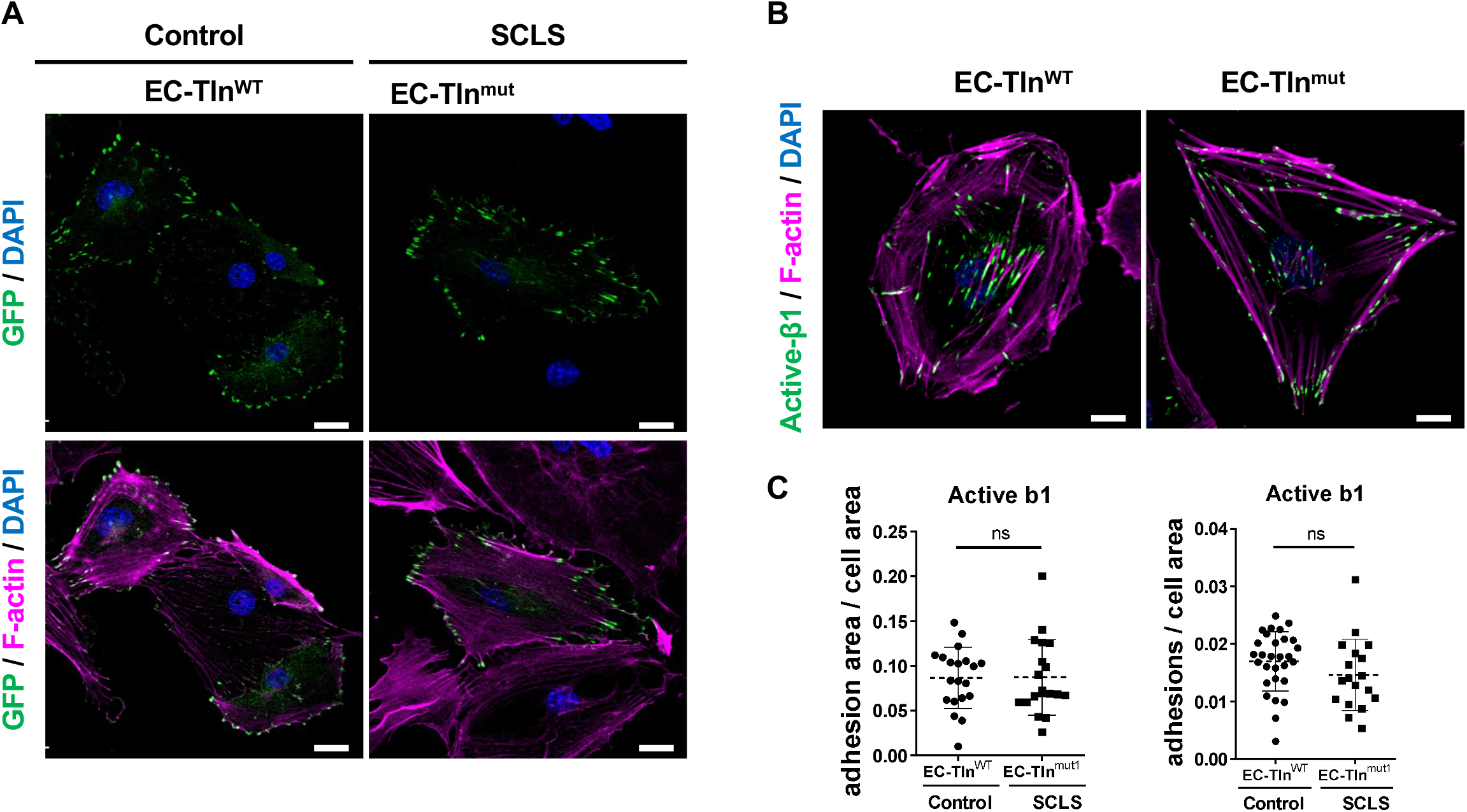
Generation of the SCLS-mimicking endothelial cell model. Representative images of primary mouse talin1 heterozygous endothelial cells transfected with full-length talin1-GFP (EC-Tln^WT^) and SCLS mutant talin1 R2510A-GFP (EC-Tln^mut^) constructs. (A) Top panels show GFP expression of the constructs (green) and phalloidin labelling F-actin (magenta), while bottom panels show only GFP expression of the constructs. (B) Immunofluorescent analysis of integrin β1 (green) and phalloidin (magenta) of control EC-Tln^WT^ and SCLS mimicking EC-Tln^mut^ heterozygous primary endothelial cells. Nuclei were stained with DAPI. Scale bar, (A) 20μm, (B) 10μm. (C) Graphs display the quantification of the cell-ECM adhesion area (μm^2^) and the number of cell-ECM adhesion sites per cell area (μm^2^) measured with IMARIS software. Data is grouped from at least 2 independent experiments, n of cells; EC-Tln^WT^: 29, EC-Tln^mut^: 19. Statistical analysis, Mann–Whitney rank-sum test. ns, no statistically significant difference.

### SCLS-mimicking talin1 mutant disrupts the integrity of endothelial adherens junctions

Using the SCLS-modelled ECs, we next set out to characterize the endothelial cell-cell junctions which we visualize using immunofluorescent analysis of VE-cadherin, a key component and regulator of adherens junctions (AJs) (Dejana and Orsenigo, 2013). Morphological examination of endothelial cell confluent monolayers formed by wild-type endothelial cells (WT) or heterozygous control endothelial cells expressing full-length talin1 protein (EC-Tln^wt^) displayed continuous and well-formed AJs. However, heterozygous SCLS EC-Tln^mut^ modelled endothelial cells revealed severely disrupted AJs compared to control heterozygous EC-Tln^wt^ or wild-type endothelial cells (Fig. 5A). The fragmentation of adherens junctions caused by the SCLS-mutation is more clearly visualized using the color-scaling, where red indicates big and continuous VE-cadherin areas (> 70μm) and blue-violet highlights small and unstable adherens junctions (< than 30μm) (Fig. 5B). Quantification of VE-cadherin staining showed decreased localization of VE-cadherin at cell-cell borders (Fig. 5C) and discontinuous adherens junctions in SCLS-Talin^mut^ endothelial monolayers compared to controls. Quantification of the number and the distribution of VE-cadherin fragments showed that SCLS EC-Tln^mut^ endothelial cells have significantly less of the large and intermediate adherens junctions and significantly more small junctional fragments compared to control cells (Fig. 5D). The decreased VE-cadherin signal in SCLS EC-Tln^mut^ AJs is not caused by alterations in total VE-cadherin expression levels (Fig. 5E) and thus indicates a functional disruption of adherens junctional dynamics as a consequence of the mutation. In support of these findings, another structural component of adherens junction, β-catenin, was similarly disrupted in SCLS EC-Tln^mut^ endothelial cells compared to control EC-Tln^wt^ (Fig.S7).

**Figure 5.**
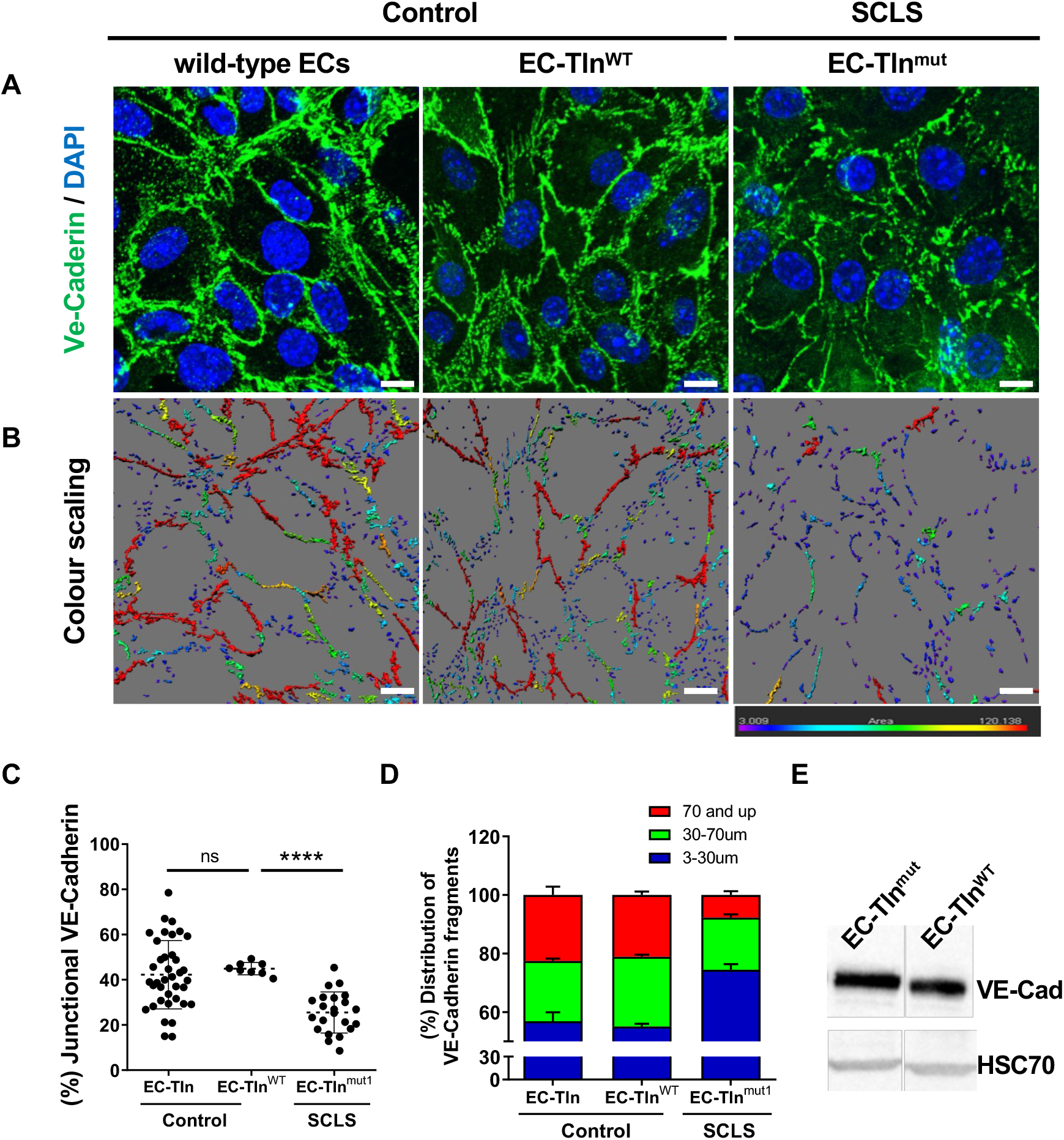
Disruption of adherens junctions in SCLS-mimicking endothelial monolayer. (A) Representative images of VE-Cadherin (green) immunostained confluent monolayers of mouse primary endothelial cells, either wild-type (WT) or heterozygous for Talin1 transfected with full-length talin1 protein (EC-Tln^wt^) or SCLS talin1 mimicking mutation R2510A (EC-Tln^mut^). Nuclei were stained with DAPI. (B) Colour scaling of the VE-cadherin staining area, whereby the red colour is the highest continuous VE-Cadherin area (>70μm), decreasing to smaller areas marked by all the different colours until it reaches the lowest measurements, which are of blue-violet colour (<30 μm), analysed by IMARIS. Scale bar, 10 μm. (C) Graph displays the quantification of the continuous junctional VE-Cadherin staining area, as represented by the percentage of VE-cadherin staining surfaces above 70μm to the total VE-cadherin staining area. (D) Graph displays the distribution of three different indexes of VE-cadherin fragments as the percentage of the total VE-Cadherin staining surfaces. In all the graphs shown, data represent the mean ± SD of at least 3 independent experiments. n of field of views analysed, wild-type ECs=30; EC-Tln^wt^= 8; EC-Tln^mut^= 22. Statistical analysis, Mann–Whitney rank-sum test. ns, no statistically significant difference, **** p<0.0001. (E) Western blot analysis of mouse primary endothelial cells transfected with the various constructs showing that the total levels of VE-cadherin remain unchanged between the control and the mutation-transfected cells. HSC70 acts as loading control.

### SCLS-mimicking mutant weakens endothelial adherens junctions by sequestering vinculin at cell-ECM adhesions

We next set out to investigate the molecular mechanism of the endothelial barrier disruption caused by the SCLS mimicking talin1 mutation. Using two different antibodies, one against the N-terminus and another against the C-terminus of the talin1 protein, we observed, as expected, robust staining of the cell-ECM adhesions at the bottom of the endothelial monolayers. However, we could not detect any co-localization of talin 1 with VE-cadherin, at AJs (Fig. 6). Therefore, we concluded that talin1 affects the endothelial barrier without being a structural component of cell-cell junctions.

**Figure 6.**
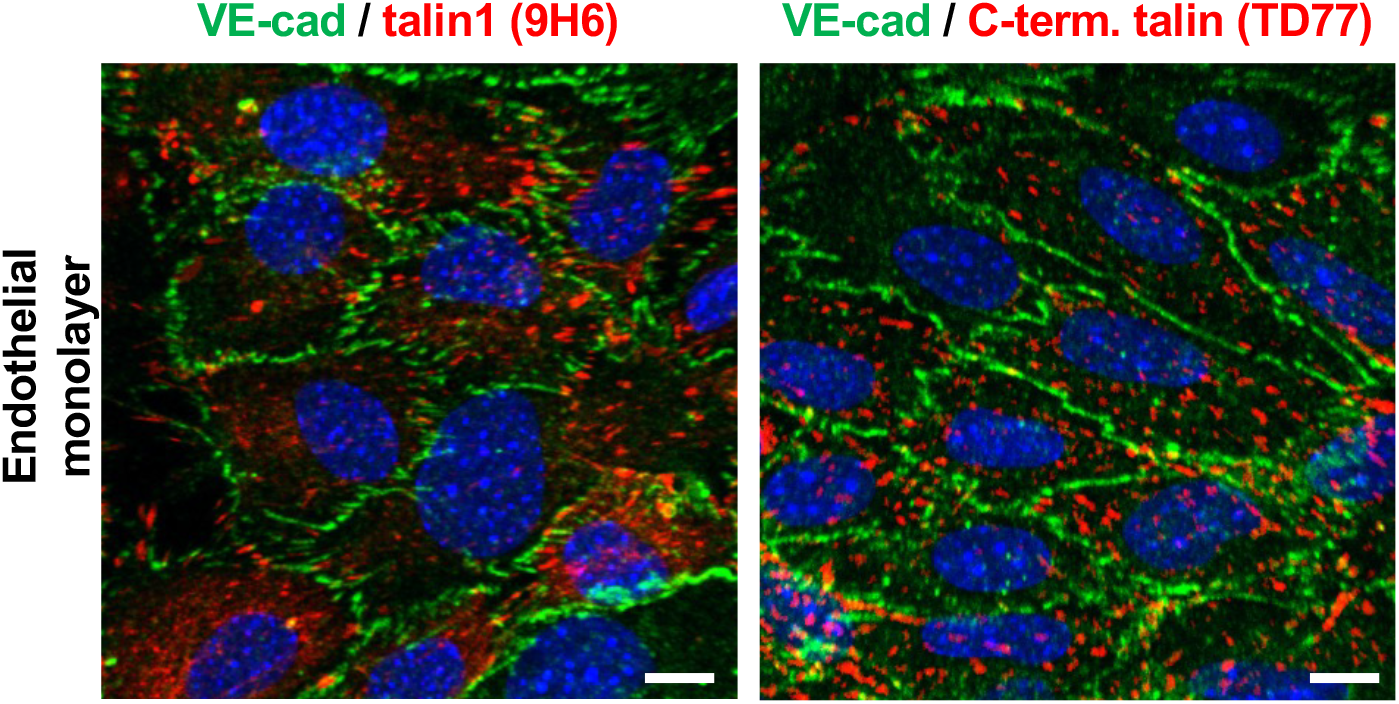
Talin 1 is not localised at VE-cadherin adherens junctions. Representative confocal images of primary endothelial cell monolayers immunostained with antibodies against VE-cadherin (green) and talin 1 97H6(red) or VE-cadherin (green) and C-terminus talin TD77 (red). DAPI depicts nuclei. Scale bars, 10 μm.

Talin has multiple binding sites for vinculin, a key adaptor that can bind to both talin and the actin cytoskeleton and thus can facilitate adhesion stability (Goult et al., 2013; Atherton et al., 2015; Gingras et al., 2005). Additionally, vinculin has been shown to be an important regulator of barrier function both in epithelial and endothelial cells (Huveneers et al., 2012; Birukova et al., 2016; Twiss et al., 2012). Therefore, we examined whether the SCLS mimicking talin1 mutation affects vinculin expression and/or co-localization. Immunofluorescent analysis revealed a significant reduction of vinculin co-localization with VE-cadherin in heterozygous SCLS-Tln^mut^ ECs compared with controls (Fig. 7A, and B). The decreased vinculin localisation at cell-cell junctions of SCLS-mimicking endothelial cells was not caused by reduced total levels of vinculin expression, as these were unchanged, as shown by western blot analysis (Fig. 7C). Interestingly, we found vinculin to be enriched in actively-remodeled cell-ECM adhesions, as demonstrated by the significantly increased co-localization of vinculin with phosphorylated pY31-paxillin (Fig. 7D, E).

**Figure 7.**
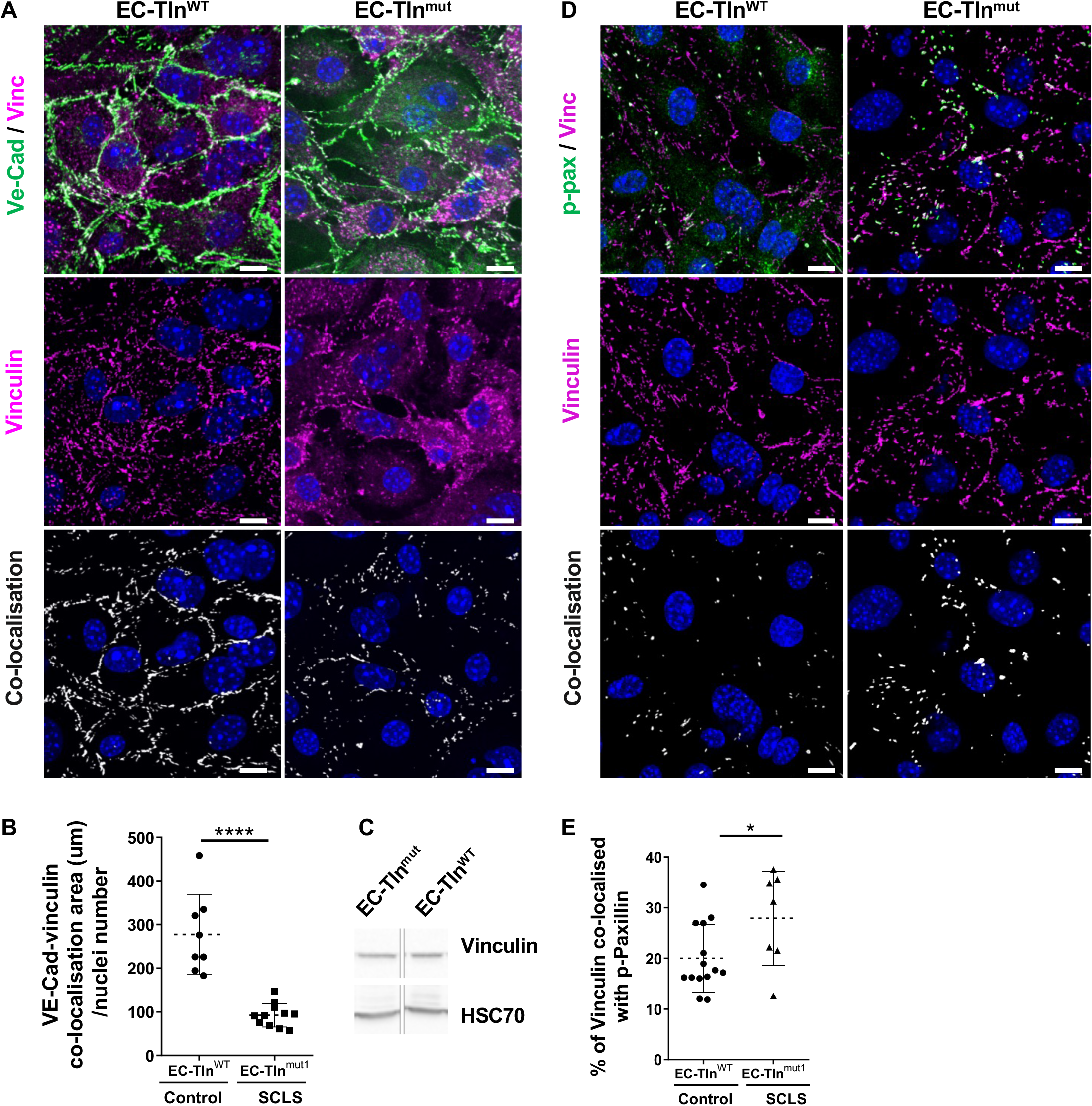
Defective vinculin-dependent stabilization of AJs in SCLS mimicking endothelial monolayers. (A) Representative images of VE-cadherin (green) and vinculin (magenta) immunostained confluent monolayers of mouse primary heterozygous talin 1 endothelial cells transfected with full-length talin 1 (EC-Tln^wt^) or the SCLS talin 1 mimicking mutation R2510A (EC-Tln^mut^). Vinculin signal alone (magenta) and the co-localisation signal of VE-cadherin/vinculin (white) are shown in the same planes (middle and bottom panel, representatively). Nuclei were stained with DAPI. (B) Graph displays the quantification of the co-localisation area of VE-cadherin and vinculin normalised to the number of nuclei present in each field of view. (C) Western blot analysis of total vinculin expression levels in EC-Tln^wt^ and EC-Tln^mut^ endothelial cells. HSC70 acted as a loading control. (D) Representative images of phosphorylated pY31-paxillin (green) and vinculin (magenta) immunostained confluent monolayers of mouse primary heterozygous talin 1 endothelial cells transfected with full-length talin 1 (EC-Tln^wt^) or the SCLS talin 1 mimicking mutation R2510A (EC-Tln^mut^). Vinculin signal alone (magenta) and the co-localisation signal of vinculin/pY31 paxillin (white) are shown in the same planes (middle and bottom panel, representatively). Nuclei were stained with DAPI. (E) Graph displays the quantification of the percentage of vinculin co-localised with pY31-paxillin at dynamically remodelled adhesion sites. In all graphs data represent the mean ± s.e.m. of at least 3 independent experiments; n of fields of view analysed for B, EC-Tln^wt^= 8; EC-Tln^mut^= 11 and n for D, EC-Tln^wt^= 14; EC-Tln^mut^= 7. Statistical analysis, Mann–Whitney rank-sum test. ****p<0.0001, **p<0.005, Scale bars, 10 μm.

Taken together, we propose a model to explain the disruption of endothelial barrier stability observed in SCLS (Fig. 8) based on alterations in vinculin localization as a result of the mutation. It has been shown that vinculin is responsible for stabilizing adherens junctions, without affecting their formation (Timmerman et al., 2015; Huveneers et al., 2012). Vinculin also can increase cell-ECM adhesion strength by binding to actin filaments (Atherton et al., 2015, 2016; Stutchbury et al., 2017). The interactions of talin and vinculin are tightly regulated and the force loading on talin is critical to this process. We propose that in SCLS disease, disturbance of talin’s actin binding capabilities results in accumulation of vinculin in the remodeling cell-ECM adhesions possibly by dysregulation of the force loading on talin. The cellular levels of vinculin are limiting and increased localisation of vinculin at cell-ECM adhesions hinders its translocation to adherens junctions. Reduction of vinculin from adherens junctions leads to less stable cell-cell junctions. Therefore, when a trigger induces endothelial permeability, adherens junctions are unable to get remodeled properly and timely, causing vascular leakage, which is the hallmark of SCLS disease.

**Figure 8.**
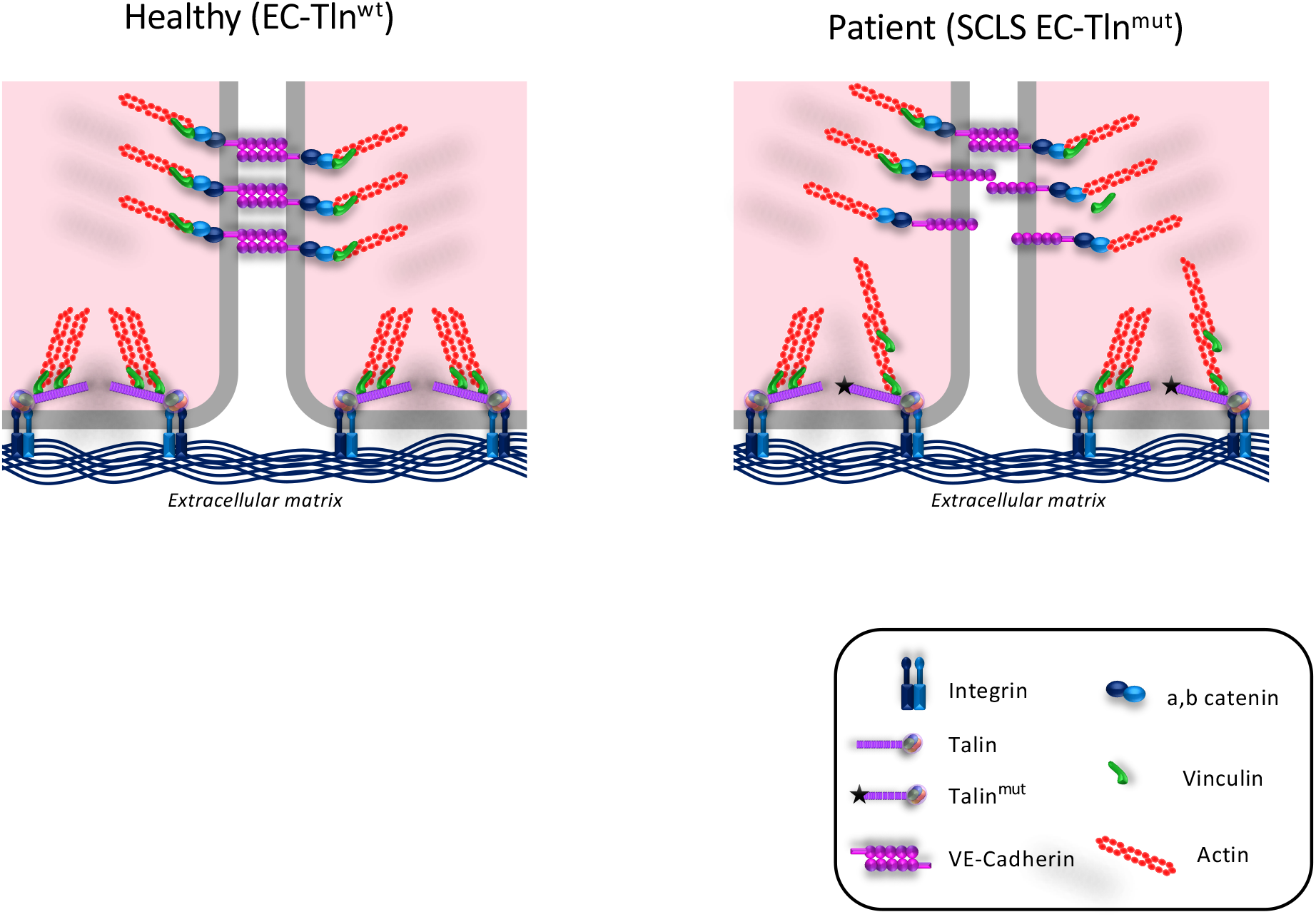
Schematic model of how SCLS mimicking talin1 mutation affects endothelial barrier function. In wild-type endothelial cells, vinculin is localised in both cell-ECM and cell-cell adhesions to regulate their dynamics. In SCLS endothelial cells with heterozygous disruption of talin1 R13 domain and predictively defective actin binding by ABS3, vinculin is sequestered at cell-ECM adhesions to stabilize actin interactions in expense to the remodelling of the adherens junctions.

## DISCUSSION

In this study we report a familial case of SCLS with three affected individuals from an extended pedigree with SCLS who each carry a rare heterozygous splice-site variant in the *TLN1* gene. This splice variant distorts the R13 talin domain and is predicted to affect the c-terminal actin binding region, ABS3. Talin1 protein is a key cell-ECM adhesion component that can activate integrins, sense and respond to mechanical forces and mediate the link to the actin cytoskeleton (Goult et al., 2018; Critchley, 2005; Critchley and Gingras, 2008). Given that the patient fibroblasts had unaltered cell-ECM adhesions and integrin functions and that talin has established roles in vascular function (Monkley et al., 2011b; Pulous et al., 2020), we modelled the effect of the SCLS-*TLN1* variant in endothelial cells heterozygous for talin1. We found that disruption of talin’s actin binding at the C-terminus disrupted adherens junction stability by sequestering vinculin to the cell-ECM adhesions. Recently, it was reported that endothelial-specific deletion of talin1 decreases VE-cadherin expression and localization at adherens junctions, causing endothelial barrier dysfunction and bleeding (Pulous et al., 2019). In contrast to a complete endothelial talin1 deletion, the heterozygous SCLS-mimicking talin1 mutation had no effect on VE-cadherin protein expression but severely disrupted its localization at cell-cell contacts, resulting in fragmented adherens junctions.

Although the trigger of the SCLS remains unknown, both inflammatory mediators and vasodilators have been found in serum from patients under attack and are believed to trigger the opening of capillary junctions and to increase permeability (Xie et al., 2012; Druey, 2014; Hsu et al., 2015). Permeability agonists, among them thrombin and histamine, induce Rho-mediated actin-myosin contractility and cause force-dependent remodeling of cell junctions, creating jacked adherens junctions (Timmerman et al., 2015; Dorland and Huveneers, 2016; Oldenburg et al., 2015). Talin has been shown to be a key mechanotransducer *in vitro* (Elosegui-Artola et al., 2016) and *in vivo* (Nikolopoulou et al., 2022) and in response to mechanical forces generated by the actomyosin contractility, talin undergoes intramolecular structural rearrangements, that reveal more binding sites for interacting partners, such as vinculin (Goult et al., 2018). Recently developed FRET probes have detailed Talin’s conformational changes in response to altered tension at cell-ECM adhesions and have shown that the C-terminal actin-binding domain (ABS3) is important for reinforcing cell-ECM adhesions (Kumar et al., 2018, 2016; Atherton et al., 2015). Our data suggests that the SCLS-*TLN1* variant disrupts talin’s ABS3 creating a dysfunctional link to actin cytoskeleton. This in turn causes vinculin accumulation at cell-ECM adhesions to reinforce binding to the actin cytoskeleton. In agreement with this, a linear correlation between mechanical forces and residence time for vinculin at cell-ECM adhesions has been reported (Dumbauld et al., 2013). The sequestration of vinculin at cell-ECM adhesions of the heterozygous SCLS mutant significantly impaired vinculin’s function at adherens junctions, thereby delaying the proper remodeling of cell-cell junctions after a trigger, impacting on the protection of the endothelial barrier function.

The intermittent presentation of SCLS is intriguing and may be due to an altered percentage of mis-spliced versus wild-type *TLN1* transcripts under various cellular conditions as a result of a currently unidentified environmental trigger. Whilst the trigger is still not established a commonly reported condition prior to a SCLS episode was dehydration and identification of the trigger and the mechanism of action might indicate pathways involved in *TLN1* gene processing. Similarly, a trigger might affect the mechanical properties of the vascular endothelium. Talin tension has been reported to regulate local actin organization and focal adhesion dynamics (Kumar et al., 2018). Our data suggests that talin’s ABS3 could be important for sensing endothelial tension and regulate vinculin localization and vascular barrier function. A potential therapeutic avenue would be to modulate and restore normal splicing by use of splice-switching antisense oligonucleotides (Havens and Hastings, 2016).

In conclusion, our data demonstrate an undescribed role for talin-C-terminal ABS3 as a regulator of endothelial barrier stability. Most importantly, these findings are directly relevant to human pathology. Understanding the molecular mechanism of SCLS and its triggers will allow for early detection of attacks and possible preventative measures and can lead to more effective focused treatments.

## MATERIALS AND METHODS

### Exome analysis

Following informed consent, exome analysis was performed on DNA extracted from whole blood of the proband and two other affected individuals and healthy control relatives (**Fig. 1A**,). Exonic sequences from DNA were enriched with the xGen Exome Research Panel IDT-V2 Kit together with the xGen Human mtDNA Research Panel v1.0 kit. Sequences were generated on a NovaSeq6000 sequencing system (Illumina, San Diego, California, USA). Read alignment and variant calling were performed with DNAnexus (Palo Alto, California, USA) using default parameters with the human genome assembly hg19 (GRCh37) as reference. Exome analysis yielded 59 million reads, with a mean coverage of 79.1 X.

### Segregation analysis

An amplicon containing the *TLN1* variant was amplified by conventional PCR of genomic DNA isolated by members of the extended family using the following primers: TLN1_DNA_F1: 5’-GGCCTAAAGCAGGGAGAGTT-3’ and TLN1_DNA_R1: 5’-GGACAGGTTGGCACTTTGAT-3’. Amplicons were analyzed by Sanger dideoxy nucleotide sequencing.

### RNA isolation and reverse-transcription PCR

RNA was isolated from peripheral blood or cultured fibroblasts of two patients and unaffected relative control by TRIzol reagent extraction or using the Qiagen RNeasy kit according to the manufacturer’s instructions. cDNA was prepared from 1 μg RNA using qScript cDNA Synthesis Kit (Quantabio or the RevertAid M-MuLV Reverse Transcriptase kit (Fermentas). The region encompassing exons 52-57 of *TLN1* was amplified by a PCR reaction using PCRBIO HS Taq Mix Red (PCR Biosystems) with primers TLN1_RNA_F1: 5’-CAGAGGACCCCACAGTCATT-3’and TLN1_RNA_R1: 5’s-TCCGAAGCATTTCTTCCTGT-3’. The resultant fragments were separated by 3% (w/v) agarose gel electrophoresis and their sequence determined by Sanger sequencing. Quantitative real-time PCR was performed using primer TlnF1 (exon 54): 5’-CAATGCACTGGACGATGGG-3’ and TlnR1 (exon 55): 5’-TGCCTCACACAGATTGTTGGT-3’ on a StepOne Plus system (Applied Biosystems) and results were analyzed with the StepOne software. The human *RPLP1* gene was used as a reference gene and was amplified with primers F: 5’-AAGCAGCCGGTGTAAATGTTGAGC-3’ and R: 5’-CATTGCAGATGAGGCTCCCAATGT-3’.

### Protein analysis

Structural modelling of the exon skipping event was done comparing the solution structure of R13 (Gingras et al., 2008) with a structural model of SCLS_R13 generated using the open-source software, ColabFold to run AlphaFold2 software (Jumper et al., 2021; Mirdita et al., 2022). Structures were visualized using PyMOL (The PyMOL Molecular Graphics System, Version 2.0 Schrödinger, LLC).

### Protein expression and purification

Synthetic genes of the murine SCLS_R13-DD (residues 2300-2541 delta2376-2395), and SCLS_R12-DD (residues 2138-2541 delta2376-2395) in the pET151 expression vector were purchased from GeneArt. The equivalent wildtype constructs were already available and described previously(Gingras et al., 2008). Recombinant His-tagged chicken vinculin domain 1 (Vd1; residues 1–258) was expressed from a pET-15b plasmid. The five constructs were each expressed in BL21(DE3) *E. coli* cells grown in Lysogeny broth (LB) with 100 μg/ml ampicillin at 37°C.

Protein expression was induced with 0.2 mM Isopropyl β-D-1-thiogalactopyranoside (IPTG) overnight at 18°C. Proteins were purified by nickel affinity chromatography, followed by anion exchange as described previously(Khan et al., 2020). R12-DD and SCLS-R12-DD were spun on their own at 50,000 x g for 20 min at 4°C. The samples were analyzed using SDS-PAGE.

### Size Exclusion Chromatography (SEC)

Size exclusion chromatography analysis of talin1 R12-DD and SCLS-R12-DD alone and in the presence of vinculin domain 1 (VD1) was performed at room temperature. Samples were run at 100 μM talin and 100 μM Vd1. The samples were loaded and ran using a Superdex 200 increase 10/300 GL.

### Circular Dichroism (CD)

Circular dichroism experiments were performed using a JASCO J-715 spectropolarimeter. Samples were 0.5 mg/ml R12-DD WT and R12-DD SCLS samples in 20 mM Tris pH 8, 150 mM NaCl. Far-UV spectra were recorded at wavelengths 200-260 nm, using 6 scans at 50 nm/min speed, 1 nm step resolution and 1 nm band width. Melting curves were collected a fixed, 222 nm wavelength, varying the temperature between 20-90°C, with 1°C step resolution and 1 nm band width.

### Primary cell cultures

Primary fibroblasts were isolated from skin samples of patients IV-3, IV-11 and healthy relatives. Cells were cultured in DMEM medium supplemented with 10% fetal calf serum and 1% Penicillin/Streptomycin and L-Glutamine. Early cell passages (P2-P8) were used for all experiments.

Mouse primary endothelial cells were isolated from lungs of *TLN1*-heterozygous or wild-type control mice as described previously (Kostourou et al., 2013). Briefly, lungs were minced, digested with 0.1% collagenase type I for 1h, passed through a 70μm pore size cell strainer and after centrifugation cells were re-suspended in mouse lung endothelial cell medium (Ham’s F-12/DMEM 1:1, 1% Penicillin/Streptomycin and L-Glutamine, 20% fetal calf serum and endothelial mitogen), and plated onto tissue-culture flasks pre-coated with a mixture of collagen I, human plasma fibronectin, and 0.1% gelatin. Endothelial cells were purified by magnetic immunosorting using intracellular adhesion molecule ICAM-2 (1:300, BD Biosciences #553325) and PECAM-1 (1:1000, BD Biosciences #553370) antibodies. Cultures were tested for endothelial purity by flow cytometric analysis using PECAM-1, VE-cadherin, and ICAM-2 antibodies. All experiments were performed in early passages (P2-P6) of pure ECs.

### Live cell imaging

To assess cell spreading, control and patient fibroblasts were seeded on coated wells of 6-well tissue culture plates and were transferred to an environmental chamber of 37°C and 5% CO_2_ coupled to a Zeiss Observer Z.1 microscope. Two different planes were chosen from each well and photos were acquired every 5 min for 72 cycles. Camera and shutter were controlled by the Zen software (Zeiss) and movies were processed using the same program.

### Primary endothelial cell transfections

Primary ECs were transfected with Talin1-wt or Talin1-R2510A constructs using the Amaxa Nucleofector kit (VP1-1001, Lonza) together with the Nucleofector electroporator (Lonza), according to the manufacturer instructions. After transfection cells were seeded in 100mm pre-coated tissue culture plates and were left to recover. Forty hours post-transfection ECs were sorted by flow cytometry (BD FACSAria II, BD Biosciences) and only the GFP positive cells were kept, which were afterwards seeded in coverslips or 8-well slides for further immunofluorescence analysis.

### Immunofluorescence analysis of Focal Adhesions

For the analysis of focal adhesion sites mouse primary ECs (either WT or transfected with the mutant talin constructs) or primary fibroblasts were seeded on 13mm round glass coverslips in 24-well culture plates at a density of 2×10^4^ cells/well. After 48 h cells were fixed with 4% PFA for 10 min at 4°C, washed three times in 1x PBS and then permeabilized with 0.3% Triton X-100/PBS for 10 min. Afterwards, cells were washed once with PBS and incubated for 1 h at room temperature in 1% BSA/PBS for blocking. Primary antibodies (anti-talin1 clone 97H6, 1:500, Bio-Rad #MCA4770; anti-talin2, 1:50, custom made; anti-paxillin, 1:200, BD Biosciences #610052; anti-phospho-Tyr 118-paxillin,1:200, Cell signalling #2541; anti-vinculin,1:400, Sigma Aldrich #V9264; anti-phospho-Tyr397-FAK,1:100, Biosource #44624G; anti-integrin-β1 clone 9EG7, BD Biosciences #550531) were added on the coverslips in 0.1% BSA/PBS blocking buffer and were incubated overnight at 4°C. The next day cells were washed three times with PBS and were incubated with the appropriate secondary antibodies (Alexa-Fluor-conjugated, 1:400, Thermo Fisher Scientific & phalloidin-633 for F-actin, 1:400, Thermo Fisher Scientific A-22284) for 1 h at room temperature in 1% BSA/PBS, where 4,6-diamidino-2-phenylindole (DAPI) was also added. After three final washes, coverslips were mounted on microscope slides with Mowiol. Images were captured using a Zeiss Observer Z.1 or a confocal SP8X WLL system (Leica Microsystems) and quantification of the focal adhesions were performed with the IMARIS software (Oxford Instruments).

### Immunofluorescence analysis of Adherence Junctions

Mouse primary endothelial cells were seeded on 8-well coated μ-slides (Ibidi) at a density of 5×10^5^ cells/well and left for 48 h to form junctions. Afterwards they were fixed using 4% PFA for 10 min at 4°C, and processed for immunofluorescence staining as described above. In the final step, cells were kept in PBS solution and were imaged directly from the 8-well slides. Antibodies used were anti-VE-Cadherin (1:100), anti-β-catenin (1:1000, Sigma #15B8), anti-talin-1 clone 97H6 (1:500, Bio-Rad #MCA4770), anti-talin1-TD77 (1:100, Merck-Millipore 05-1144), anti-vinculin (1:400, Sigma Aldrich #V9264), anti-phospho-paxillin (1:200, Cell signalling #2541) and phalloidin-633 for F-actin (1:400, Thermo Fischer Scientific A-22284). All secondary antibodies were Alexa-Fluor-conjugated (Thermo Fisher Scientific) and were used at a 1:400 dilution. Images were acquired using a confocal SP8X WLL system (Leica Microsystems) and analyzed with the IMARIS software (Oxford Instruments). For quantification of the junctional integrity, the VE-Cadherin or β-catenin signal was translated into surface objects, which were further colour-coded and categorised according to their sizes through the vantage plot function.

### Western Blot analysis

For protein levels analysis mouse primary endothelial cells or human primary fibroblasts were either lysed directly or were serum starved overnight (in DMEM with 1% FCS) and then serum-stimulated (in DMEM with 10% FCS) for 10 min before lysis. Lysis buffer consisted of 3% SDS, 60 μM sucrose, 65 mM Tris-HCl pH=6.8 supplemented with protease and phosphatase inhibitors (Roche). Protein concentration was determined using the BSA Protein Assay kit (Thermo Scientific) and 30-60μg of protein from each sample were loaded onto 10-12% polyacrylamide gels. After electrophoresis proteins were transferred to PVDF membranes, blocked using 5% milk in Tris-buffered saline with 0.1% Tween-20 (TBS-T) and incubated overnight with the primary antibodies diluted in 3% BSA/TBS-T at 4 °C. The next day blots were washed three times with TBS-T and then incubated with the relevant HRP-conjugated antibodies 5% milk/TBS-T for 1 h at room temperature. After another three washes chemiluminescence was detected using the Luminata Crescendo Western HRP substrate and a ChemiDoc XRS+ imaging system (BioRad). Densitometry readings were acquired using the Image Lab software. The antibodies used included anti-phospho-Ser473-AKT, anti-AKT, anti-phospho-p44/42 MAPK, anti-p44/42 MAPK, anti-phospho-Tyr118-paxillin, anti-phospho-Tyr397-FAK (1:1000, Cell Signaling #4060, #4685, #9101, #9102, #2541, Thermo #44624G); anti-talin-1 clone 97H6 (1:10000, Bio-Rad #MCA4770); (anti-talin2, 1:200, custom made); anti-VE-Cadherin (1:1000, BD Biosciences 555289); anti-integrin-β1 (1:1000, Abcam #ab3167); anti-vinculin (1:1000, Sigma Aldrich #V9264), anti-HSC70 (1:5000, Santa Cruz Biotechnology #sc7298); the appropriate HRP-conjugated anti-mouse, anti-rabbit or anti-rat antibodies (1:5000, Jackson Immunoresearch #115-035-008, #115-035-046, #115-036-03) were used as secondaries.

### Flow cytometry

For β1-integrin analysis control and patient fibroblasts were harvested by trypsinization, washed and then treated with either PBS alone, or PBS supplemented with 1 mM MnCl_2_ for 20 min. After two washes cells were incubated with the primary antibodies (total integrin-β1 Abcam #ab3167, integrin-β1 clone 9EG7, BD Biosciences #550531) diluted 1:50 in FACS buffer (2% FCS in PBS with 0.05% Sodium azide) for 45 min at 4°C. After another two washes, cells were incubated with the secondary antibodies (Alexa-Fluor anti-rat or anti-mouse 488, Thermo Fischer Scientific #A-11017, A-11006) in FACS buffer for 30 min on ice, washed again and re-suspended in PBS to be analysed on a BD FACS CantoII instrument with the FlowJo software.

### Platelet adhesion tests

Platelet adhesion was measured by the cone and plate(let) analyzer (CPA) as previously described (Savion and Varon, 2006). Briefly, whole blood was placed in polystyrene plates and subjected to a shear rate of 1,800 s-1 using a rotating Teflon cone. The plates were then thoroughly washed with distilled water, stained and analyzed with an inverted microscope connected to an image analysis system. Two parameters of platelet adhesion were evaluated: percent of platelet surface coverage (SC, %) and the average size (AS, μm^2^) of the platelet aggregates bound to the surface.

### Statistical analysis

The GraphPad Prism software (v 6.0) was used for statistical analysis. All data are presented as mean ± SD for at least 2-3 independent experiments, as stated in the corresponding figure legends. Statistical significance was determined by unpaired Student’s *t*-test or the nonparametric Mann-Whitney test when needed, where a *p* value of <0.05 was considered statistically important.

## Data Availability

All data produced in the present work are contained in the manuscript

## Author contributions

N. Elephant reported the proband IV-3, collected clinical data, analyzed genomic data, and wrote the manuscript, P. Nikolopoulou performed FACs experiments, conducted all the biochemical and cellular analysis of patient fibroblasts and wrote the paper, V. Papadaki generated the SCLS-modelled endothelial cells and analyzed the adherens junctions, D. Oz-Levi performed exome sequencing and analyzed gemonic data, G. Rouni isolated primary mouse endothelial cells and participated in cell assays and data analysis, C. Arapatzi performed molecular studies of patient fibroblasts and quantitative RT-PCR experiments, S. Yanovsky-Dagan performed molecular studies on patient-derived samples, A.R. Cowell and W. Edwards performed biochemical and structural analysis, V. Meiner analyzed exome sequencing data, R. Sion-Sarid reported the proband IV-11 and collected clinical data, V. Vainstein collected clinical data, S. Grammenoudi, performed FACs sorting and provided advice on FACs analysis, D. Lancet oversaw genetic analysis and data interpretation, B. Goult performed biochemical and structural analysis and interpret data, wrote and revised the manuscript, T. Harel oversaw clinical and genetic data interpretation, and revised the manuscript, V. Kostourou conceptualize the study and designed experiments, analyze and interpret data, acquired funding, wrote and revised the manuscript.

## Acknowledgements

The authors wish to thank the families for their participation in this study. This work was supported by the Hellenic Foundation for Research and Innovation (H.F.R.I.) under the “First Call for H.F.R.I. Research Projects to support Faculty members and Researchers and the procurement of high-cost research equipment grant” (Project Number: HFRI-FM17-2644) and the Operational Programme "Human Resources Development, Education and Lifelong Learning" in the context of the project “Strengthening Human Resources Research Potential via Doctorate Research” (MIS-5000432), implemented by the State Scholarships Foundation (IKY). B.T. Goult was funded by Biotechnology and Biological Sciences Research Council grants (BB/N007336/1 and BB/S007245/1), and Human Frontier Science Program grant (RGP00001/2016).

## Data availability

The ClinVar accession number for the DNA variant data reported in this manuscript is SCV002525869.

## Compliance with ethical standards

Informed consent was obtained in accordance with IRB-approved protocol 0306-10-HMO.

---

**Figure S1.**
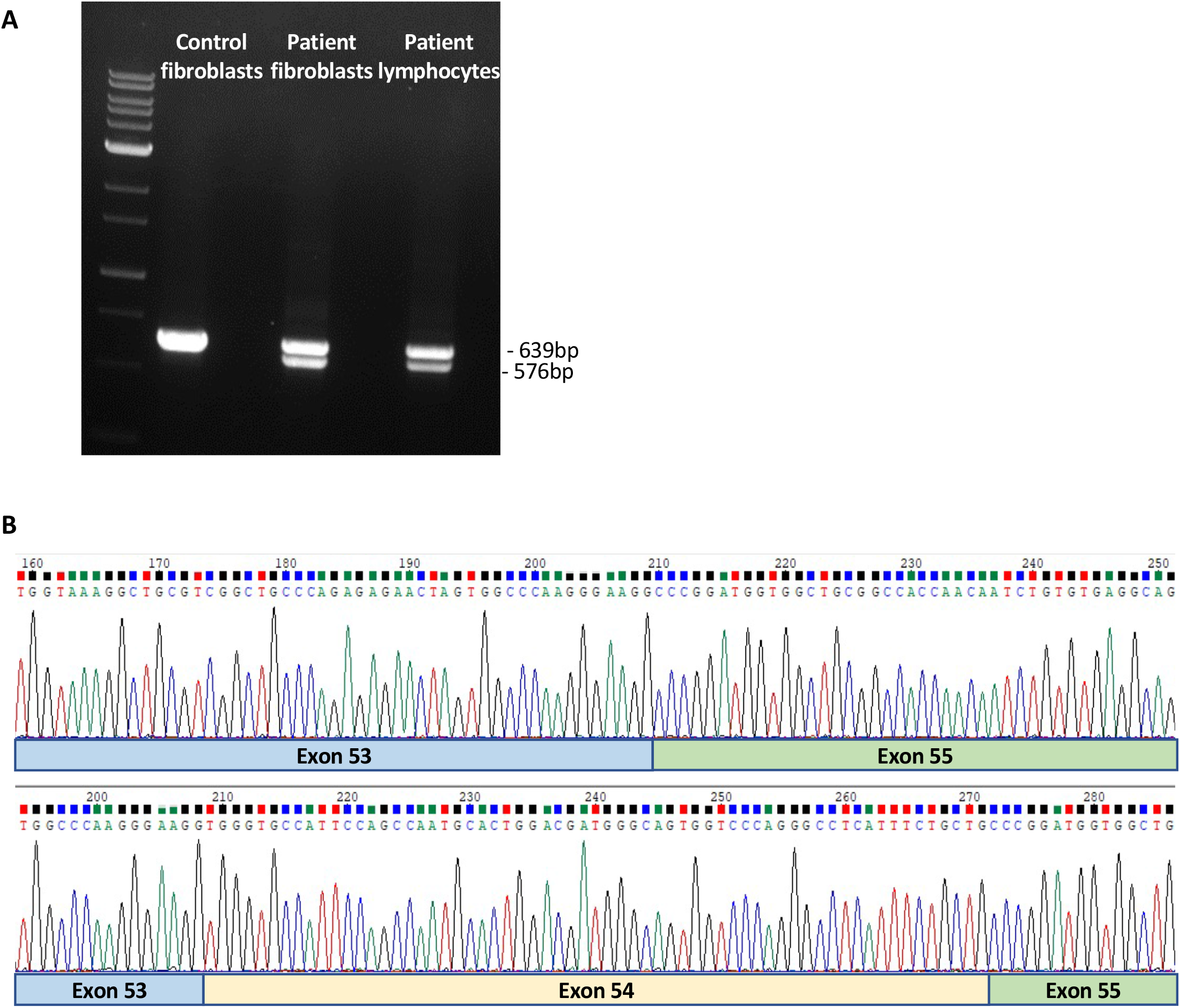
Reverse-transcription of SCLS-*TLN1* RNA demonstrate the deletion of exon54 in patient fibroblasts and lymphocytes. (A) Agarose gel of cDNA from patient and control samples, demonstrating a lower molecular weight band in patient fibroblasts and lymphocytes (from peripheral blood) as compared to control fibroblasts, indicating mis-splicing. wild-type cDNA: 639 bp; mutant cDNA with skipped exon 54: 576 bp. (B) Sanger sequencing at the cDNA level. Upper panel represents the sequence of the mutant allele following gel extraction, showing in-frame skipping of exon 54. Lower panel shows the wild-type sequence. Data shown are means ±SD from 3 independent experiments in triplicates.

**Figure S2.**
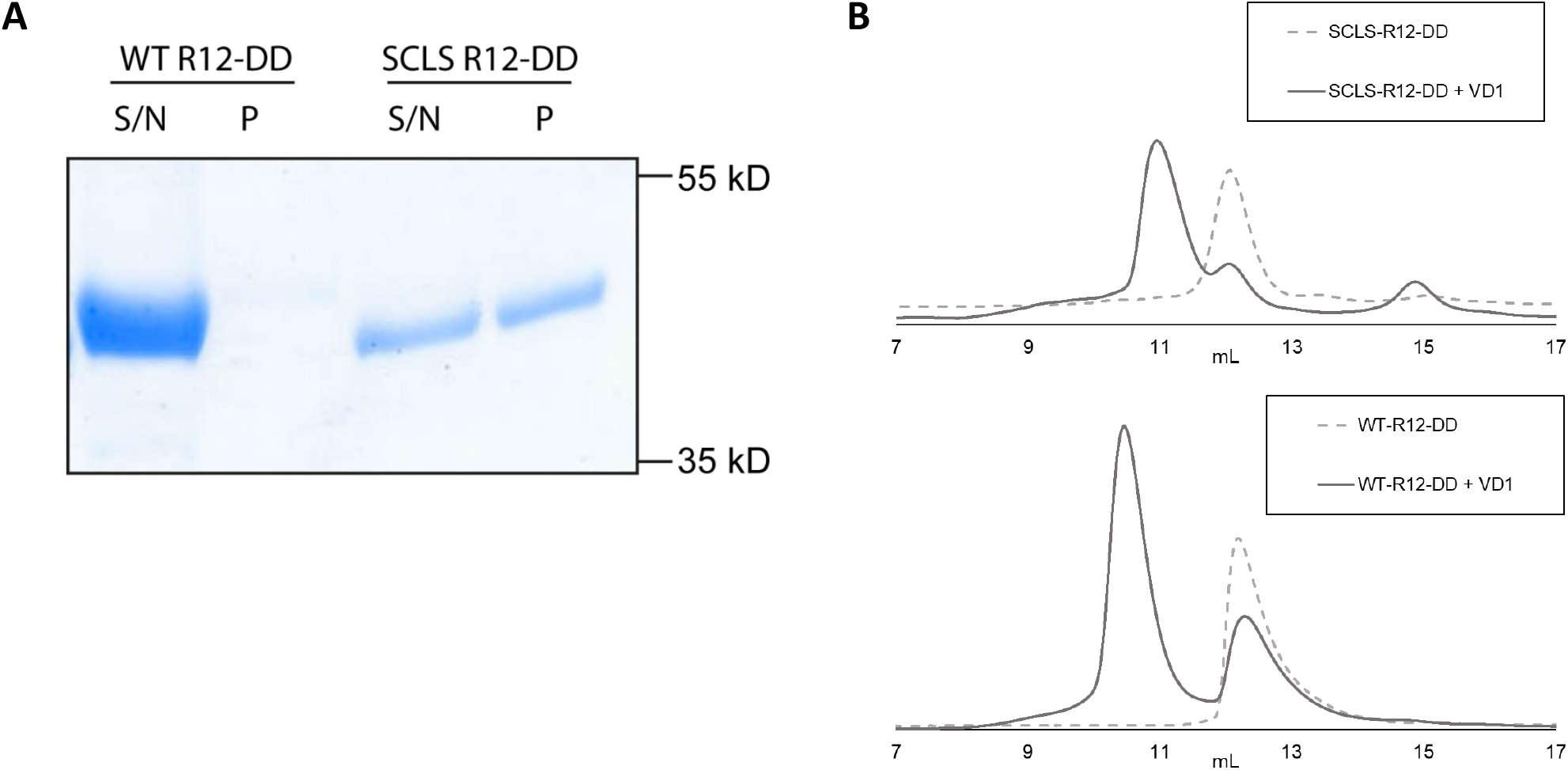
Biochemical analysis of the SCLS-R12-DD domain. (A) Control experiment of the high-speed centrifugation for the actin co-sedimentation experiment. WT-R12-DD and SCLS-R12-DD were spun alone at 50,000 x g. WT is seen in the supernatant (S/N) whereas the SCLS-R12-DD was present in the S/N and the pellet (P). (B) Size exclusion chromatography (SEC) analysis showing 100 μM SCLS-R12-DD (top) and WT-R12-DD (bottom) alone and titrated with 100 μM VD1.

**Figure S3.**
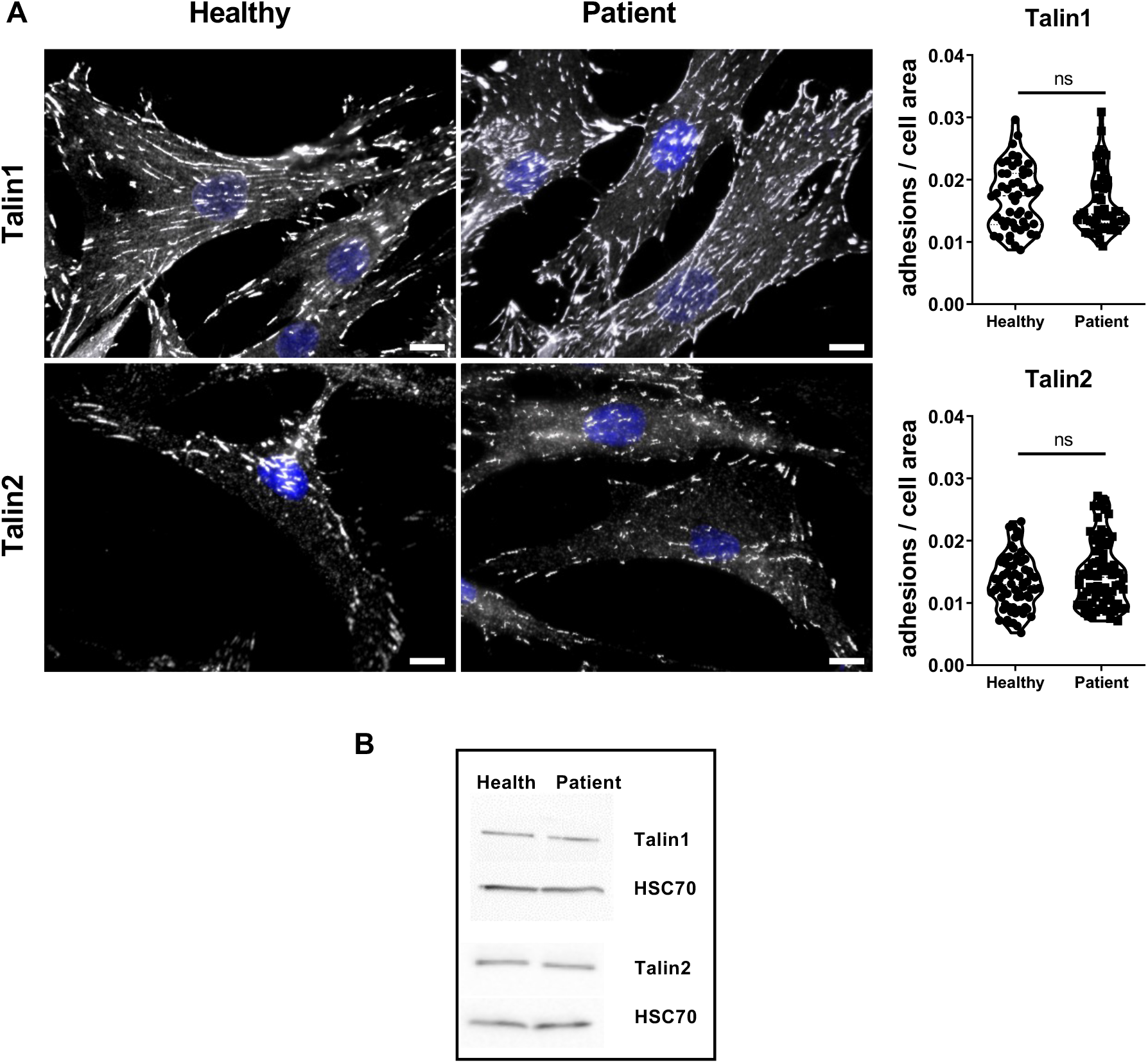
Talin1 and talin2 protein expression and localisation is not altered in SCLS patient fibroblasts. (A) Immunofluorescence staining of healthy donor and SCLS patient fibroblasts with antibodies against talin1 (upper panel) and talin2 (bottom panel). Scale bar 15μm. Violin plots display the number of cell-ECM adhesions per cell area (μm^2^) as quantified with IMARIS software. Data is grouped from 3 independent experiments, n of cells; talin1: Healthy=50, Patient=50; talin-2: Healthy=66, Patient=78. Statistical analysis, Mann–Whitney rank-sum test. ns, no statistically significant difference. (B) Western blot analysis of the talin1 and talin2 expression in lysates from healthy and patient fibroblast. HSC70 acts as the loading control.

**Figure S4.**
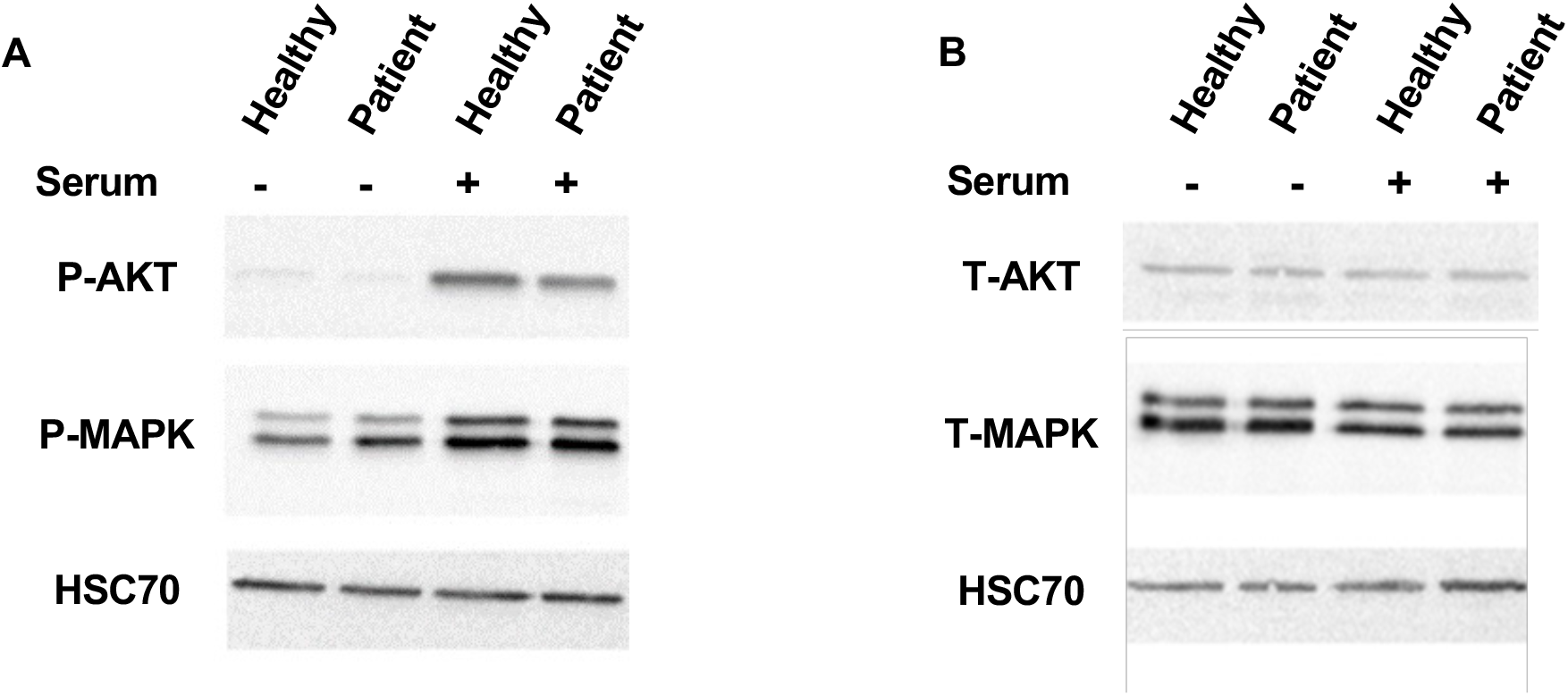
The SCLS-TLN1 variant does not affect signal transduction in human fibroblasts. Western blots of (A) pS473 AKT, pY42/44 MAPK (pMARK) and (B) total AKT (tAKT), total MAPK (tMAPK), in lysates of SCLS patient and healthy donor fibroblasts before and after serum stimulation for 10 mins. HSC70 was used as loading control.

**Figure S5.**
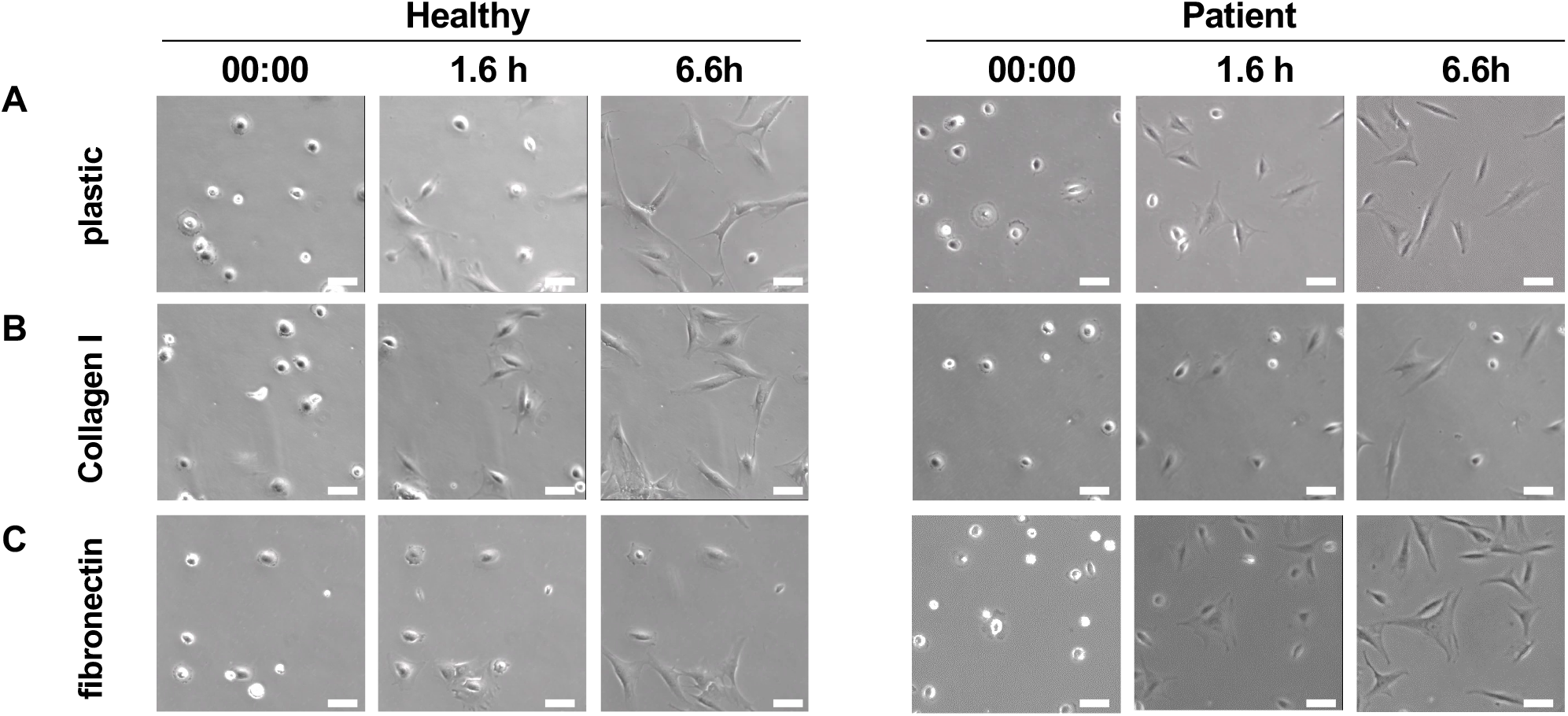
Cell adhesion and spreading is unaltered in SCLS patient and healthy fibroblasts. Selected frames from different time points of time-lapse imaging showing cell adhesion and spreading on plastic, collagen I and fibronectin of healthy donor and SCLS patient fibroblasts.

**Figure S6.**
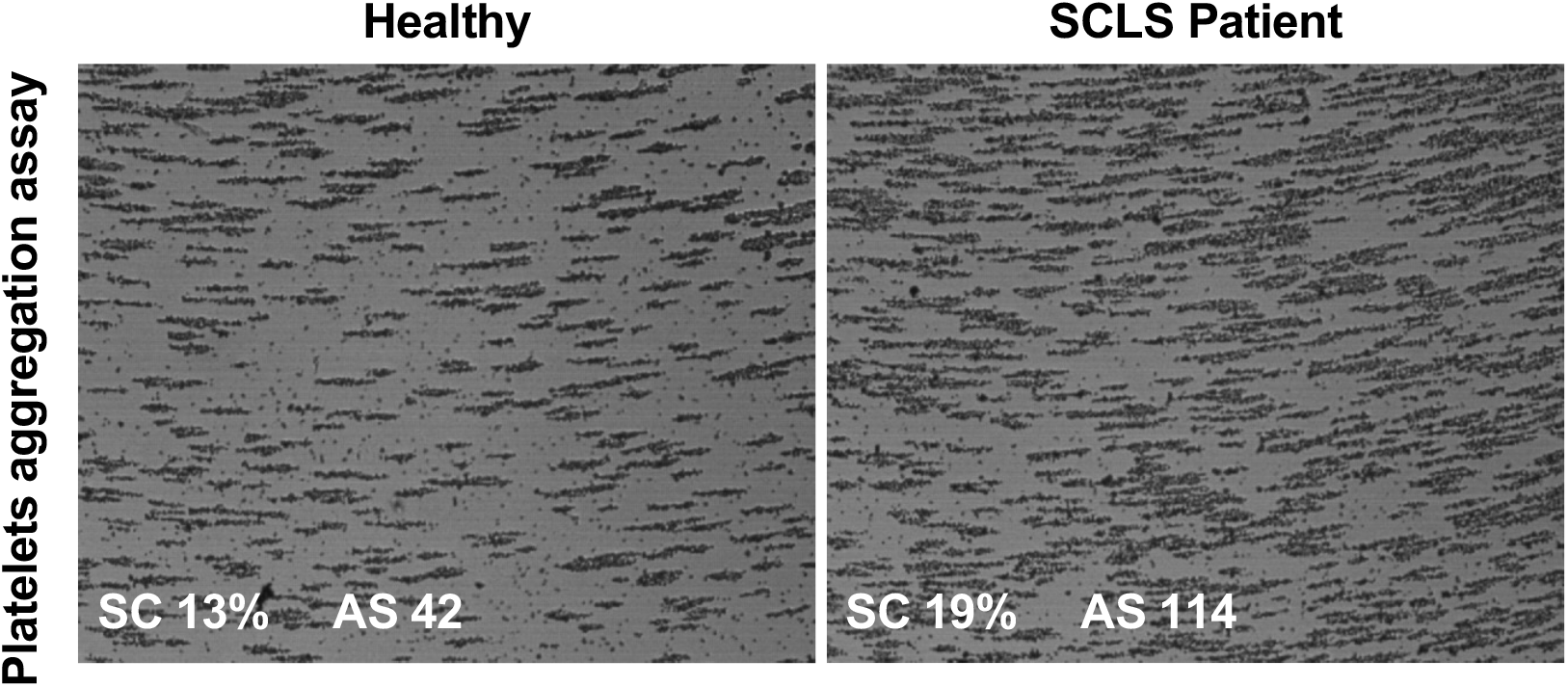
Platelets are unaffected in SCLS patient. Platelet’s aggregation assay. Testing the blood sample from both the patient IV-5 and the healthy relative donor results in aggregates formation on the well surface. SC; surface coverage. AS; average size.

**Figure S7.**
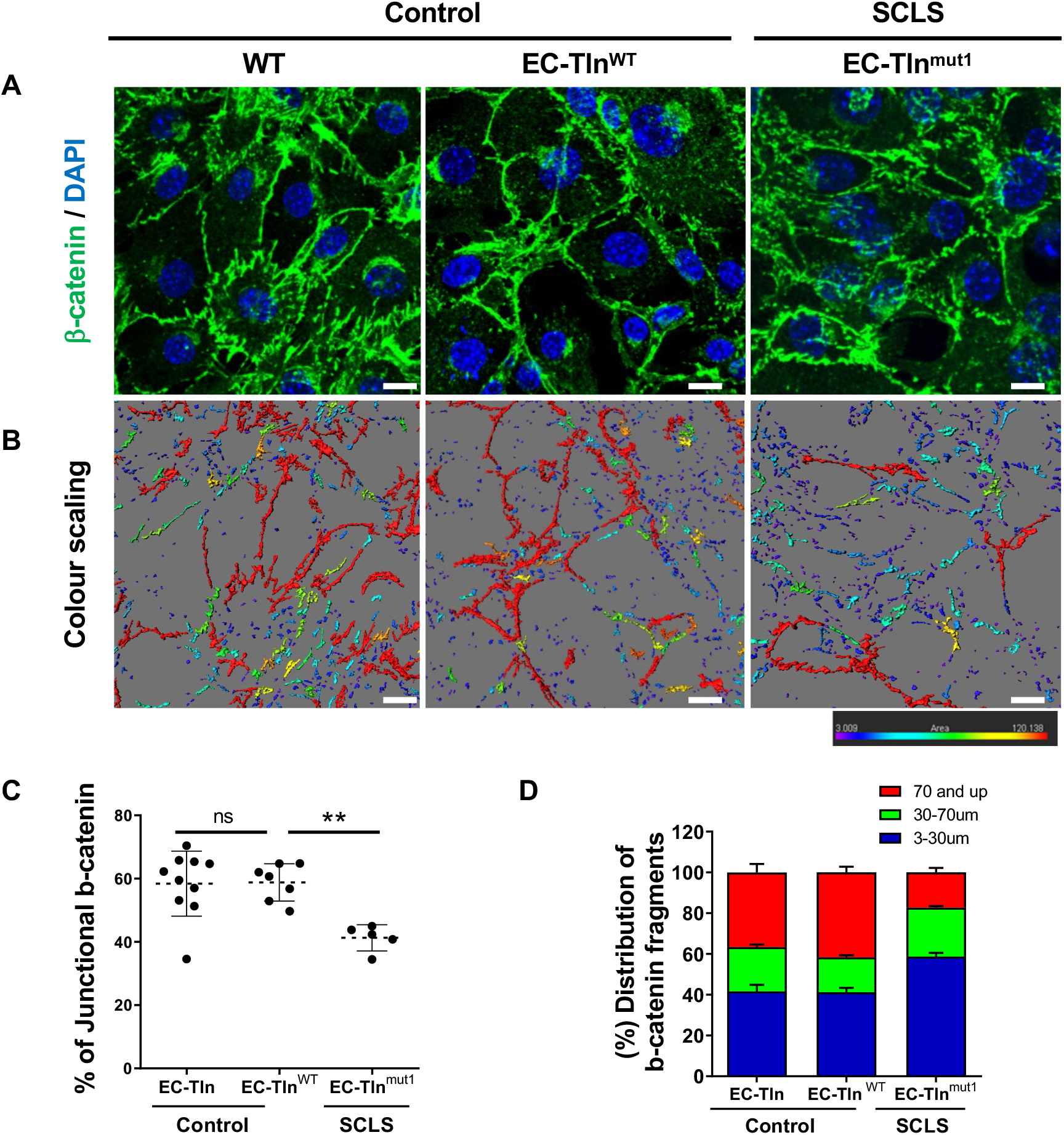
Disrupted adherens junctions visualised using b-catenin staining in SCLS mimicking endothelial cells. (A) Representative images of β-catenin (green) stained confluent monolayers of mouse primary endothelial cells, either wild-type (WT) or heterozygous for Talin1 transfected with full-length talin 1 protein (EC-Tln^wt^) or SCLS talin1 mimicking mutation R2510A (EC-Tln^mut^). Nuclei were stained with DAPI. (B) Colour-scaling of the β-catenin staining area, whereby the red colour is the highest continuous β-catenin area (>70μm), decreasing to smaller areas marked by all the different colours until it reaches the lowest measurements, which are of blue-violet colour (<30 μm). Scale bars, 10 μm. (C) Graph displays the quantification of the continuous junctional β-catenin staining area, as represented by the percentage of b-catenin staining surfaces above 70μm to the total β-catenin staining area. (D) Graph displays the distribution of three different indexes of β-catenin fragments as the percentage of the total β-catenin staining surfaces. Data represent the mean ±SD of two experiments, n of field of view analysed EC-Tln^wt^=7; EC-Tln^mut^= 5. Statistical analysis, Mann–Whitney rank-sum test. ns, no statistically significant difference, ** p<0.005

## Video 1 Legend

Movie of combined time-lapse imaging showing cell adhesion and spreading on different substrates (plastic, collagen I and fibronectin) of healthy donor and SCLS patient fibroblasts. Scale bar 30μm.

